# Vertical topography in EEG microstates: Physiology or artifact manifestation?

**DOI:** 10.1101/2024.07.29.24311153

**Authors:** Tomáš Jordánek, Martin Lamoš, Radek Mareček

**Affiliations:** First Department of Neurology, St. Anne’s University Hospital and Faculty of Medicine, Masaryk University, Kamenice 5, Brno, 62500, Czech Republic; Brain and Mind Research Program, CEITEC, Masaryk University, Kamenice 5, Brno, 62500, Czech Republic

**Keywords:** simultaneous EEG/fMRI, EEG, microstates, microstate analysis, artifacts

## Abstract

The analysis of EEG microstates is a useful method for exploring large-scale networks and brain dynamics. In addition to the often-reported microstates, or so-called “canonical microstates”, another topography has been reported in the literature – topography with a prominent straight line separating positive and negative values that extends from the nasion to the inion (vertical topography - VT). This topography was also revealed in our simultaneous EEG/fMRI and shielded cabin EEG data collected from 77 participants.

Following analyses based on human and phantom data, we conclude that VT partially reflects artifacts caused by unspecified movements of the EEG cap and its metallic components. Our conclusion is supported by evaluation of spatiotemporal characteristics of VT estimated from EEG acquired under various conditions, especially by significant correlation between the framewise displacement (obtained from human EEG/fMRI) and the temporal characteristics of VT. We recommend cautious interpretation of VT when revealed in the data. Its very presence as a resulting topography may affect the spatiotemporal parameters of the other microstates and distorts the shapes of the other topographies.

**Key points:**

- Vertical microstate topography (VT) is often present in EEG/fMRI data.
- In EEG/fMRI, VT mainly represents artifacts.
- Analysis of EEG microstates is disrupted by the presence of VT.

## 1. Introduction

Dietrich Lehmann and his colleagues (Lehmann et al., 1987) observed that scalp EEG topography remains stable for a short time (80–120 ms) and then quickly changes into another stable topography. These identified stable topographies are called microstates and are considered to be the building blocks of consciousness and mental processes, sometimes called “atoms of thought” (Khanna et al., 2015; Lehmann et al., 1998). The results of many studies have shown that just a few topographies usually explain over 70% of variance; these topographies are frequently the same across studies, they are labeled as A–D, and they are often referred to as “canonical microstates” (Michel and Koenig, 2018).

Several studies attempted to find a link among resting-state networks (RSNs) derived from functional magnetic resonance imaging (fMRI) data and microstates with the expectation that a particular microstate may be generated by a particular RSN (Britz et al., 2010; Khanna et al., 2015; Musso et al., 2010; Xu et al., 2020; Yuan et al., 2012). The conclusions were rather inconsistent with only partial overlaps. The inconsistency could have been caused by slight differences in methodology, variable numbers of resulting microstates, or the numbers of participants. Certainly, the quality of the preprocessing of the EEG data acquired in the MR environment plays a key role.

The two most prominent artifacts – gradient artifacts (GA) and pulse artifacts (PA) – are commonly handled by the average artifact subtraction (AAS) algorithm (Allen et al., 2000, 1998) or by its modifications (Acharjee et al., 2015; Bullock et al., 2021; Warbrick, 2022). The remnants of these artifacts and other sources may be suppressed by independent component analysis (ICA). The MR system itself generates vibrations that induce artifacts of known origin and therefore correctable, e.g. helium cooling pump (Rothlübbers et al., 2015) and MR bore ventilation (Nierhaus et al., 2013). Other non-neural source of artifacts, for example due to the signal induced by the interaction of AAS algorithms applied for GA and subsequent PA reduction (Steyrl and Müller-Putz, 2018) or with an elusive origin or source, such as the interaction of moving metallic parts of the EEG system with the magnetic field or changing fields, are not corrected because of the lack of appropriate methods. All these sources of artificial EEG signal components certainly bias microstate analysis.

Among the studies that use EEG microstates analysis, a significant number can be found that report another topography in addition to the canonical microstates – a left-right topography with a vertical line in the middle – the vertical topography (VT). Some studies found VT at the subject level to be one of the dominant topographies (D’Croz-Baron et al., 2021; Xu et al., 2020). Other studies (Agrawal et al., 2022; Bréchet et al., 2019; Custo et al., 2017; Rajkumar et al., 2021; Schwab et al., 2015; Thiele et al., 2023; Zappasodi et al., 2019) have even reported VT as one of the resulting group microstates. Interestingly, it seems that the presence of VT does not depend on which EEG or MR system was used, nor on the specific type of preprocessing or microstate analysis pipeline.

VT is reported more often in simultaneous EEG/fMRI studies (Agrawal et al., 2022; Rajkumar et al., 2021; Schwab et al., 2015; Zappasodi et al., 2019) than in studies that use EEG data acquired in a shielded room without the presence of MR environment (Bréchet et al., 2019; Custo et al., 2017; Thiele et al., 2023). Although we cannot rule out that VT may have a physiological origin under certain circumstances, we hypothesize that VT is to some extent a reflection of EEG artifacts caused mainly by various movements of the EEG system during data acquisition that are even amplified in the MR environment. To confirm this hypothesis, we analyzed characteristics of VT in our human data and prepared a few phantom measurements that were mainly aimed at finding the origin of VT in the data. We show the distorting effect VT has on microstate analyses.

## 2. Materials and methods

### 2.1 Human data

#### 2.1.1 Participants

The dataset consists of 77 subjects (39 M, 38 F; mean age 31.4 ± 7.4 years) with no signs of neurological or psychiatric disorders. All of them signed informed consent forms before the measurements were taken. EEG recordings in a human shielded cabin (HSC) and simultaneous EEG/fMRI (human magnetic resonance – HMR) were obtained from each participant, 20 minutes of resting-state data with closed eyes was recorded. The EEG cap stayed placed on the subject’s head for both recordings to maintain the same electrode positions.

#### 2.1.2 EEG recording and preprocessing

The EEG data obtained both in the shielded cabin and during simultaneous EEG/fMRI were recorded by the MR-compatible HD-EEG system EGI GES 400MR with a 256-channel cap (256-channel HydroCel Geodesic Sensor Net; Electrical Geodesics, Inc., Eugene, Oregon). The sampling frequency was 1 kHz, and all electrodes were referenced to Cz electrode. Impedances of all electrodes were kept under 50 kΩ.

GA and PA in HMR EEG data were reduced with the AAS algorithm (Allen et al., 2000, 1998) in the BrainVision Analyzer (BrainVision Analyzer, Version 2.1.1, Brain Products GmbH, Gilching, Germany). All subsequent preprocessing steps were the same for both recording environments (shielded cabin and MR) and were done in Matlab (MATLAB version: 9.8.0.1359463 (R2020a) Update 1, Natick, Massachusetts: MathWorks Inc.). Face and neckline electrodes were discarded because of a higher inclination to muscle artifacts and conductivity failures. The final EEG data comprised signals from 204 electrodes. Data were manually inspected, and bad channels and epochs with artifacts were marked and excluded from ICA and microstate analysis. The filtering was performed by an 8^th^ order Butterworth bandpass filter of 1–40 Hz. Eye blinks, oculomotor artifacts, and electrocardiogram (mainly in HSC EEG) were suppressed by ICA. On average, two independent components were removed from HMR EEG (eye blinks, eye movements) and four components (eye blinks, eye movements and heart activity) were removed from HSC EEG. Selection of ICA components was rather conservative – only components that clearly represent above mentioned artifacts were removed. Signals from bad channels were interpolated with a 3D spherical spline from neighboring channels. At the end, data was downsampled to 125 Hz.

#### 2.1.3 Microstate analysis

Microstate analysis was done with freely available Cartool software (version 4.09 7492) (Brunet et al., 2011). The approach of extracting topographies only at sites of global field power (GFP) peaks was chosen (Bréchet et al., 2019; Pascual-Marqui et al., 1995). The GFP data were spatially filtered (Michel and Brunet, 2019) and clustered first at the subject level with a modified K-means algorithm (Pascual-Marqui et al., 1995). The optimal number of clusters was selected based on Metacriterion, which consists of six criteria implemented in Cartool. The topographies resulting from the best clustering of each subject were clustered in a group-level analysis, again with modified K-means (200 runs).

Each time-point of the data was labeled based on a spatial correlation with group maps. The time-points that showed low correlation (<0.5) with all maps were not labeled and were omitted from subsequent analyses. Segments shorter than 3 time-points were replaced with a neighboring map. Spatiotemporal parameters were then computed for each fitted topographical map: global explained variance (GEV), mean duration, time coverage, and occurrence. The microstate template explorer toolbox was used to quantify the similarity between our microstate topographies and the four canonical microstate topographies clustered from 42 studies (Koenig et al., 2024).

#### 2.1.4 fMRI scanning and preprocessing

The fMRI data were acquired using the Siemens Prisma 3T MR machine with a 64-channel head coil. A gradient-echo EPI sequence was used: TR=650 ms; TE=15/33/52 ms; FA=30°; voxel size=3×3×3 mmm; in-plane matrix size=64×64; 48 axial slices. Each dataset was realigned by SPM12 to get the time-series of six movement parameters (translations along and rotations around the *x*, *y*, and *z* axes).

#### 2.1.5 Evaluation of subject movement

To quantify the movement, a framewise displacement (FD) metric based on movement parameters was used (Patel et al., 2014; Power et al., 2012). The mean FD value for each subject was then correlated with spatiotemporal parameters of each microstate to test for the potential relationship between microstate manifestation in the data and amount of subject movement during EEG/fMRI data acquisition. Pearson correlation was applied, and the level of statistical significance was set to p<0.05, FDR corrected.

### 2.2 Phantom data

To test our hypothesis that VT reflects movements, a series of phantom measurements were prepared. A spherical phantom for MRI filled with NiSO_4_.6H_2_O was used. The surface of the phantom was covered with a fabric soaked in the solution for EEG caps to ensure similar impedances after mounting the EEG cap on the phantom as in the human experiments. Phantom testing had three independent measurements, so the process of mounting the EEG cap to the phantom was done three times.

#### 2.2.1 Phantom dataset A

About five minutes of EEG/fMRI (phantom magnetic resonance – PMR) with exactly the same fMRI sequence and MR environment settings as for human subjects were recorded. The phantom was then moved to the shielded EEG cabin (the EEG cap position remained same) and ten minutes of EEG were recorded (phantom shielded cabin – PSC).

#### 2.2.2 Phantom dataset B

The second measurement with the phantom (phantom conditions of measurement – PCM) was set to investigate the influence of particular aspects of the MR environment on the EEG data, and specifically on VT presence. We acquired the data under eight different conditions, each lasting five minutes, with various combinations: helium cooling pump on/off, MR bore ventilation on/off, and fMRI protocol running/not running (i.e., MR gradients on/off). Another two measurements were carried out with a completely shut down MR system (i.e., standby mode) with the phantom inside or outside of the bore. All conditions are described in Table 1.

**Table 1:**
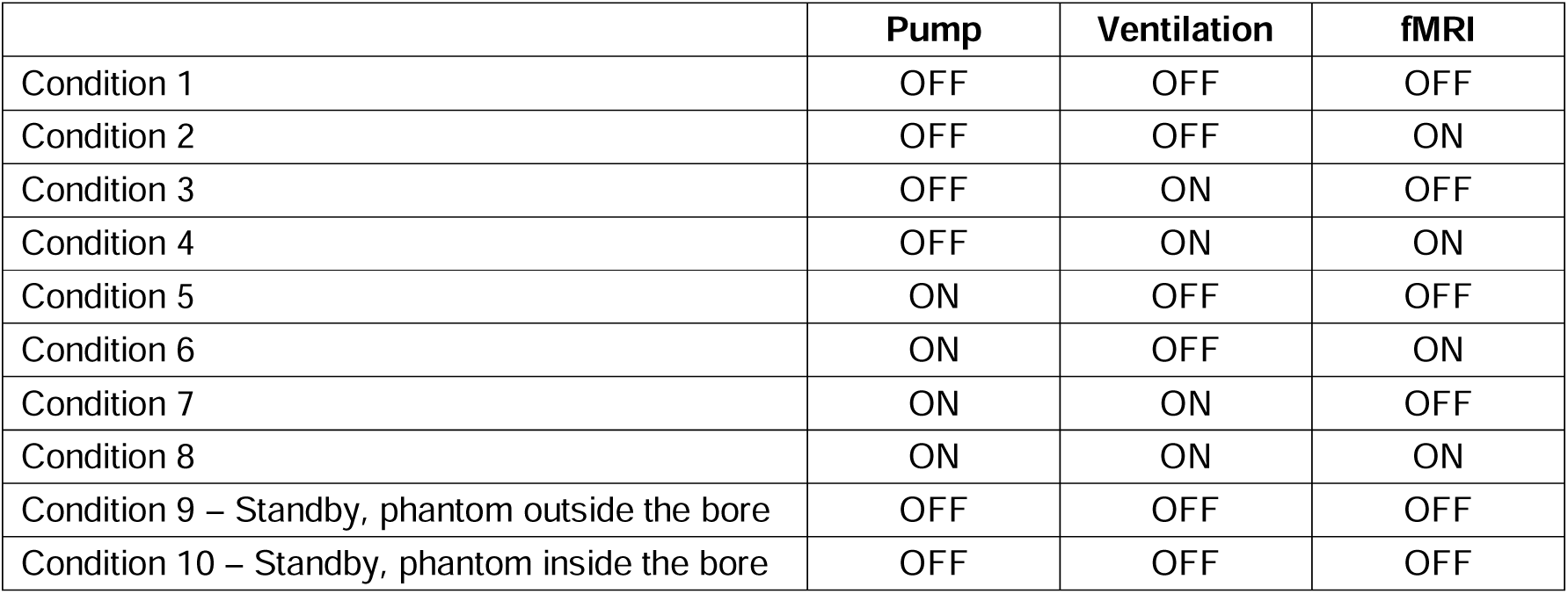
Overview of setup for each measured condition.

|  | Pump | Ventilation | fMRI |
| --- | --- | --- | --- |
| Condition 1 | OFF | OFF | OFF |
| Condition 2 | OFF | OFF | ON |
| Condition 3 | OFF | ON | OFF |
| Condition 4 | OFF | ON | ON |
| Condition 5 | ON | OFF | OFF |
| Condition 6 | ON | OFF | ON |
| Condition 7 | ON | ON | OFF |
| Condition 8 | ON | ON | ON |
| Condition 9 – Standby, phantom outside the bore | OFF | OFF | OFF |
| Condition 10 – Standby, phantom inside the bore | OFF | OFF | OFF |

#### 2.2.3 Phantom dataset C

Third phantom dataset (phantom field lowering - PFL) captures a unique opportunity to measure an EEG during the lowering static magnetic field B_0_ to zero in the course of MR machine upgrade. Except standard preparation of phantom described above we covered whole phantom into a foil to reduce drying out of electrodes and thus increasing an impedances. During the whole measurement, impedances were around 12 kOhms and increased by 0.62 ± 0.32 kOhms in average, mainly in first half of experiment (Suppl. figure 1). In the beginning, around 15 minutes were recorded with stable 3T field, then during 2 hours the field almost linearly decreased to zero, in the end, around 10 minutes were recorded without field present. In the middle of the decrease, measurement of EEG was interrupted to check for the impedances of electrodes. During the measurement, a technician several times stepped inside MR room and interacted with MR machine physically, which caused some vibrations of the whole system. He did not directly interact with phantom. During the measurement, values of current in the main coil were collected and marked to the EEG signal. As a dependence of strength of magnetic field on current in the main coil is linear (Eq. 1), values of current were directly used to evaluate changes of magnetic field.

**Eq. 1**: A formula from Siemens datasheet used for conversion between current in the main coil and strength of field *B*; *I_a_* is actual current, *I_f_*is final current for 3T, *g* is gyromagnetic coefficient of proton.

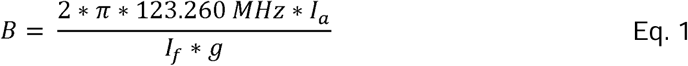

#### 2.2.4 Metrics for evaluation of relationship between VT and strength of static magnetic field

As GEV characterizes explained variance by microstate or a stable state across the EEG signal, it was used for evaluation of amount of VT that changed over time. A window approach was used and, in each 10 seconds window, a GEV value was computed according the computation in Cartool (GEV is computed only for TFs identified as VT).

Additionally to GEV, ratio between unlabeled and labeled TFs in each 10 second window was calculated. A TF is not labeled if a spatial correlation of its 2D topographic map and any resulting stable state topography is lower than 0.5. This metric helps to evaluate how the stable states are able to represent EEG data while the static magnetic field decreases.

As a third metric, entropy of EEG signal was calculated to examine a changing amount of spatial structure in the data while the static magnetic field lowered. Similarly, as in case of GEV, window approach with 10 seconds long window was used. Entropy was calculated with Matlab function “entropy” which is used for greyscale images, as entropy was calculated for whole window matrix (204 channels x 10 000 TFs) to capture spatial characteristics of EEG signal.

The preprocessing pipeline of all phantom data was similar to the human datasets (HSC, HMR). Cardioballistogram correction and ICA were not used because it was not possible for physiological artifacts to be present in the phantom data. Finally, a microstate analysis pipeline with the same settings as for human data was applied. In the PCM dataset, each condition was clustered independently to obtain representative topographies of stable states. These were then clustered again in a group-level analysis to obtain the overall representative topographies.

## 3. Results

### 3.1 Human data

Although the HSC EEG data were optimally described by four microstates (mean number of individual microstates 5.06 ± 0.73), we decided to present clustering to 6 clusters for direct comparison of the amount of VT in each dataset (6 clusters were enough to reveal VT in all the datasets), the optimal clustering for each dataset (if 6 clusters were not optimal) is in the Supplementary material. Less than 1% of the data was unlabeled and 85.7 % of variability was explained. Microstate HSC 1 was present the most often (GEV 24.2% and time coverage 30.7%). In the segmentation into four clusters (Suppl. figure 2), VT was not present. It was present in segmentations to six clusters (Figure 2, HSC 4). For details about spatiotemporal parameters see the Suppl. tables 1, 2.

The HMR EEG data were optimally described by six microstates (mean number of individual microstates 4.86 ± 0.88; Figure 2 HMR), which explained 85.8% of variance. Approximately 4% of the data was unlabeled. The most present map was HMR 6 (GEV 18.0% and time coverage 28.5%). The second most dominant map was HMR 1; it was VT (GEV 13.5% and time coverage 18.1%). For details about spatiotemporal parameters see the Suppl. table 3.

The amount of subject movement significantly positively correlated with the parameters of microstate HMR 1 (Table 2, Figure 1): the GEV and time coverage (p<0.05 FDR), and mean duration and occurrence (p<0.05 uncorrected). The correlation between FD and spatiotemporal parameters of all other microstates was negative.

**Table 2:**
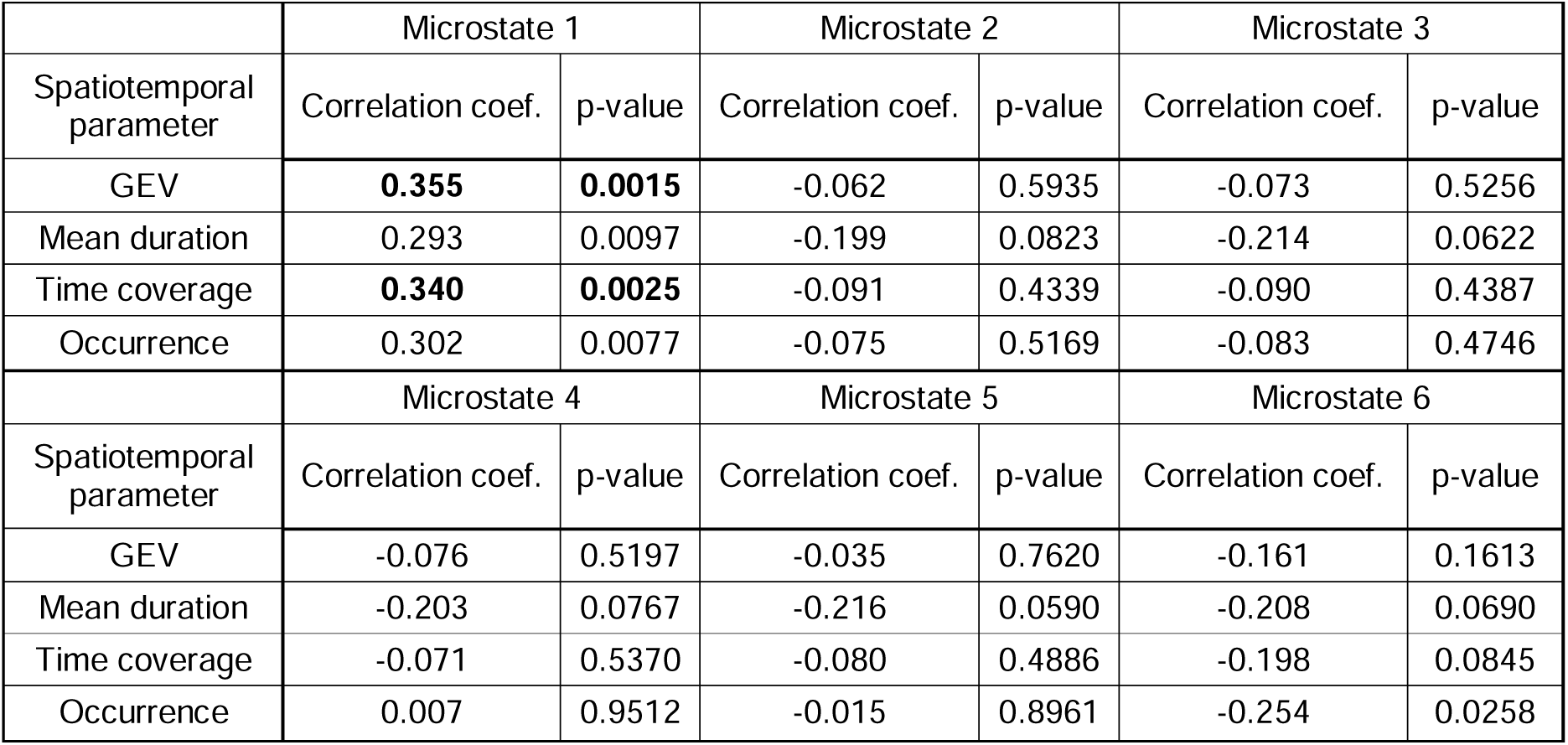
Correlation coefficients of each spatiotemporal parameter of microstates with mean FD; bold font is used for significant values after FDR correction.

|  | Microstate 1 |  | Microstate 2 |  | Microstate 3 |  |
| --- | --- | --- | --- | --- | --- | --- |
| Spatiotemporal parameter | Correlation coef. | p-value | Correlation coef. | p-value | Correlation coef. | p-value |
| GEV | **0.355** | **0.0015** | -0.062 | 0.5935 | -0.073 | 0.5256 |
| Mean duration | 0.293 | 0.0097 | -0.199 | 0.0823 | -0.214 | 0.0622 |
| Time coverage | **0.340** | **0.0025** | -0.091 | 0.4339 | -0.090 | 0.4387 |
| Occurrence | 0.302 | 0.0077 | -0.075 | 0.5169 | -0.083 | 0.4746 |
|  | Microstate 4 |  | Microstate 5 |  | Microstate 6 |  |
| Spatiotemporal parameter | Correlation coef. | p-value | Correlation coef. | p-value | Correlation coef. | p-value |
| GEV | -0.076 | 0.5197 | -0.035 | 0.7620 | -0.161 | 0.1613 |
| Mean duration | -0.203 | 0.0767 | -0.216 | 0.0590 | -0.208 | 0.0690 |
| Time coverage | -0.071 | 0.5370 | -0.080 | 0.4886 | -0.198 | 0.0845 |
| Occurrence | 0.007 | 0.9512 | -0.015 | 0.8961 | -0.254 | 0.0258 |

**Figure 1:**
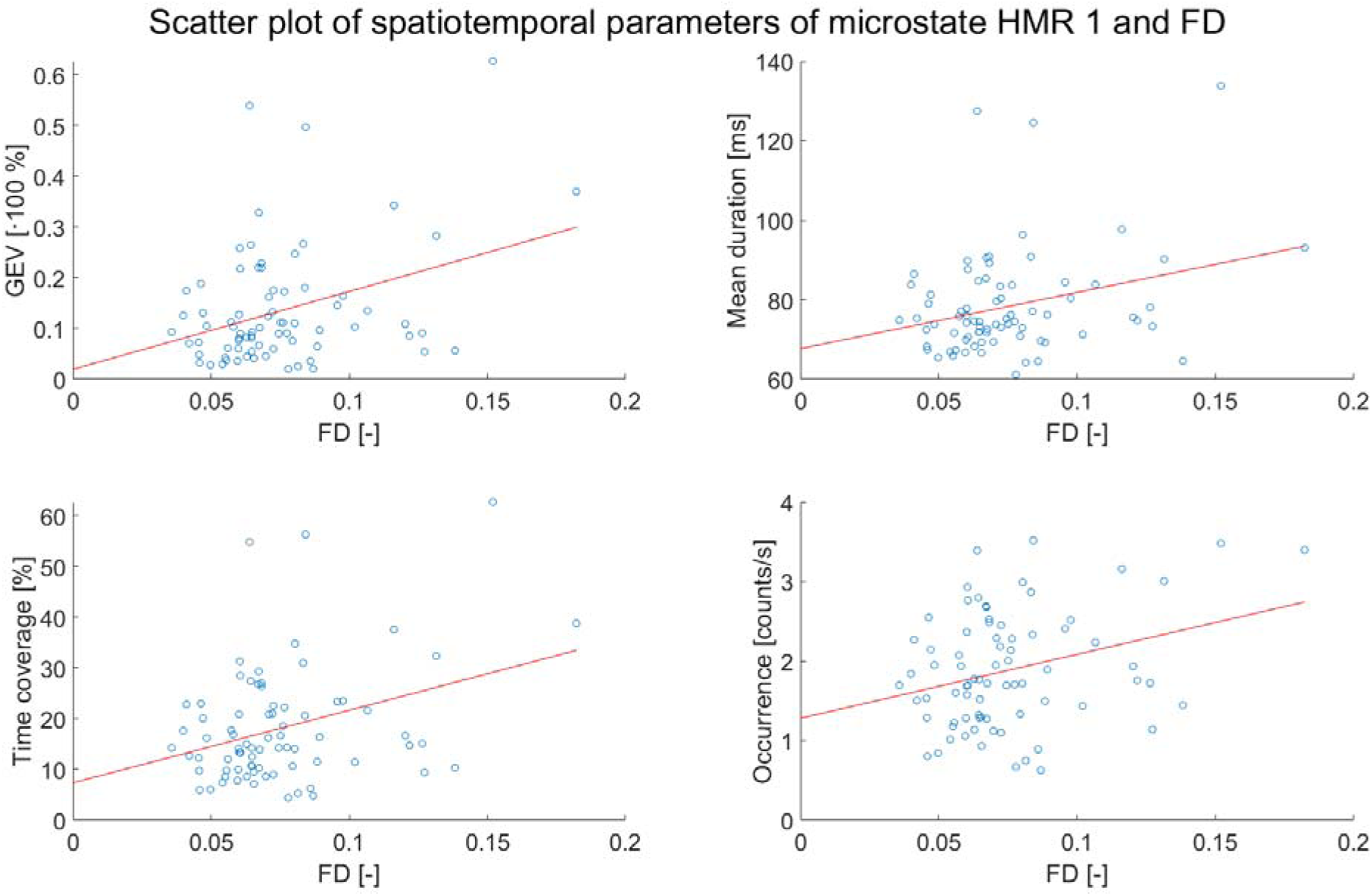
Visualization of relationship between parameter FD and spatiotemporal parameters of microstate HMR 1.

The VT was revealed in 53% of subjects for HSC data (HSC 4 in 41 out of 77 subjects) and in 92% of subjects for HMR data (HMR 1 in 71 out of 77), details are listed in Supplementary material (Suppl. figures 6-9). The median (inter-quartile range) of correlation coefficient (considering absolute value to account for polarity ambiguity) was 0.924 (0.898 – 0.942) for HMR and was significantly higher when compared to 0.952 (0.920 – 0.972) for HSC (p=0.0003, Suppl. figure 10).

### 3.2 Phantom data

The results of the phantom measurements are presented in two subsections according to the measuring sessions. The spatiotemporal parameters of the stable states are in the Suppl. tables 5–7.

#### 3.2.1 Phantom dataset A

An analysis of PMR EEG revealed eight stable states as optimum (Suppl. figure 4). We present 6 stable states (Figure 2 PMR) explaining 96.8% of GEV. Four of the stable topographies (PMR 2, 3, 4, 6) had a strong vertical line in the middle, the most dominant PMR 4 explained 55.4% of GEV and covered 59.2% of the signal. 99.9% of the data was labeled.

**Figure 2:**
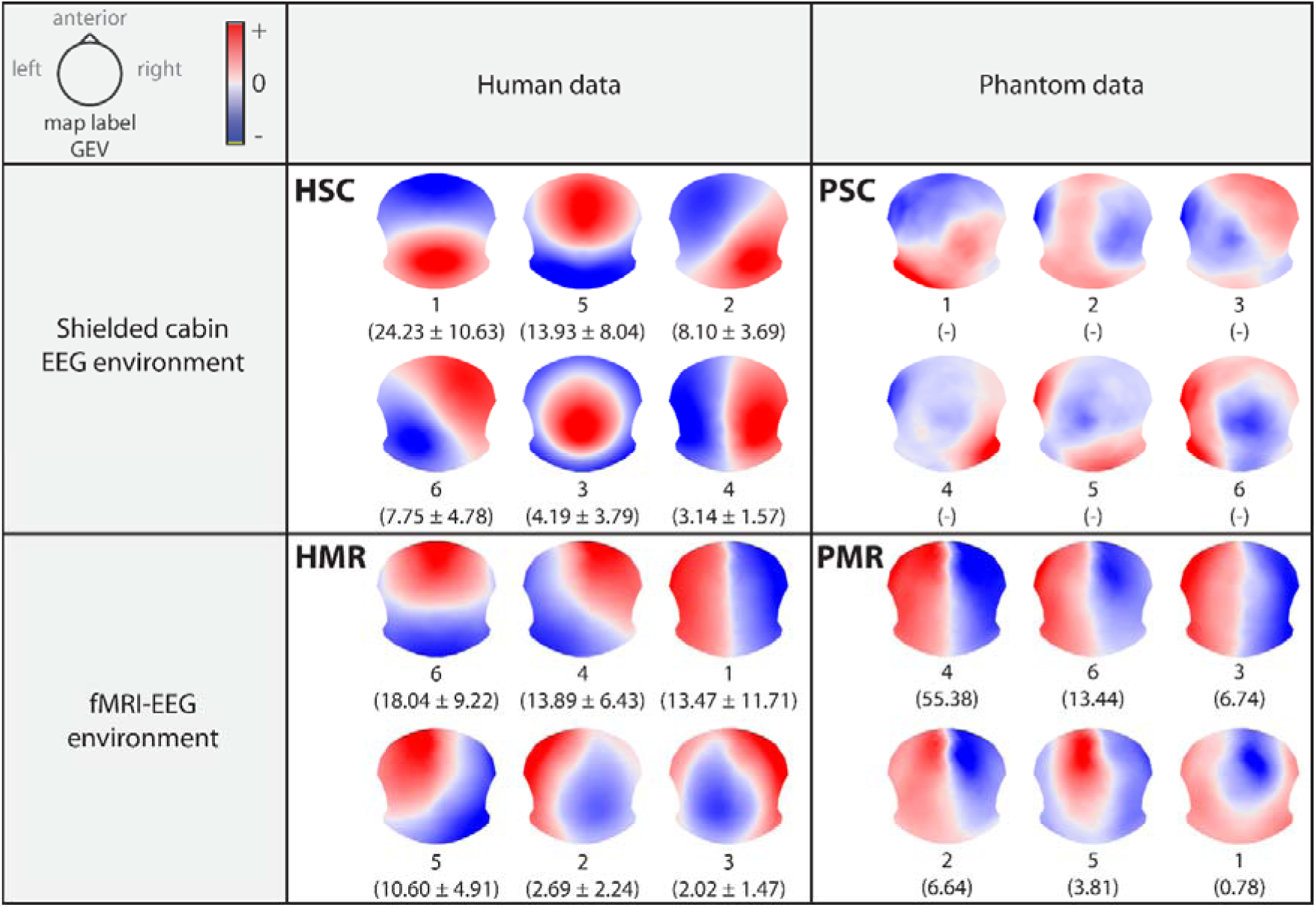
Resulting microstate topographies for human data and stable topographies for phantom data sorted by GEV value in descending order. HSC – human shielded cabin EEG data; HMR – human EEG/fMRI data; PSC – phantom shielded cabin EEG data (phantom dataset A); PMR – phantom EEG/fMRI data with the same protocol used with human subjects (phantom dataset A)

An analysis of PSC EEG data revealed seven stable states as optimum (Suppl. figure 5), again we present 6 stable states (Figure 2 PSC). Segmentations were made up to ten clusters and analyzed to check for VT presence. No segmentation revealed any signs of VT. It is important to note that 99% of the EEG is unlabeled, so the spatiotemporal parameters of stable states are not quantified here. For other segmentations, please see Suppl. figure 5.

#### 3.2.2 Phantom dataset B

Four to six maps were the optimum for the ten conditions; at least one was always VT. The group-level analysis revealed seven stable states (Figure 3) explaining 94.7% of GEV. Less than 0.1% of the data was unlabeled in each condition except for condition 9, where 4% of the data was unlabeled.

**Figure 3:**
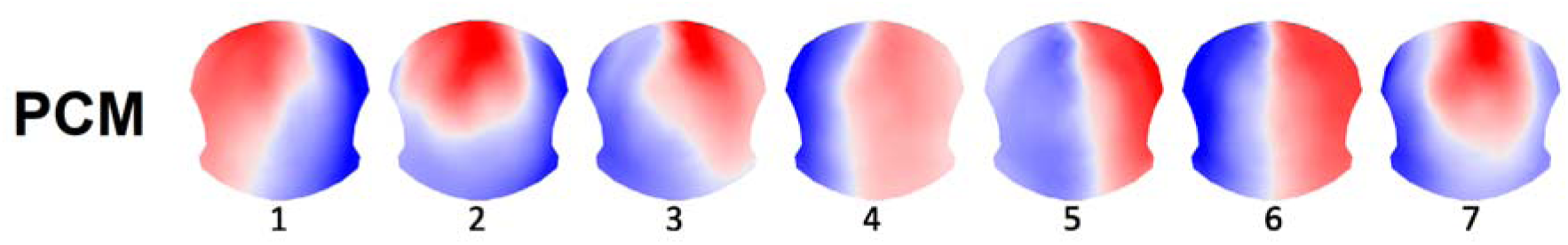
Resulting stable topographies for dataset PCM – phantom EEG/fMRI with 10 conditions of measurement

The values of GEV and time coverage respectively were summed for all three VT-like topographies (PCM 4, 5, and 6) to facilitate comparisons among conditions. Figure 4 shows that VT is the most dominant topography in conditions 1 to 8. During the MR standby mode, the presence of VT was slightly decreased when the phantom was inside the bore and notably decreased when it was outside of the bore.

**Figure 4:**
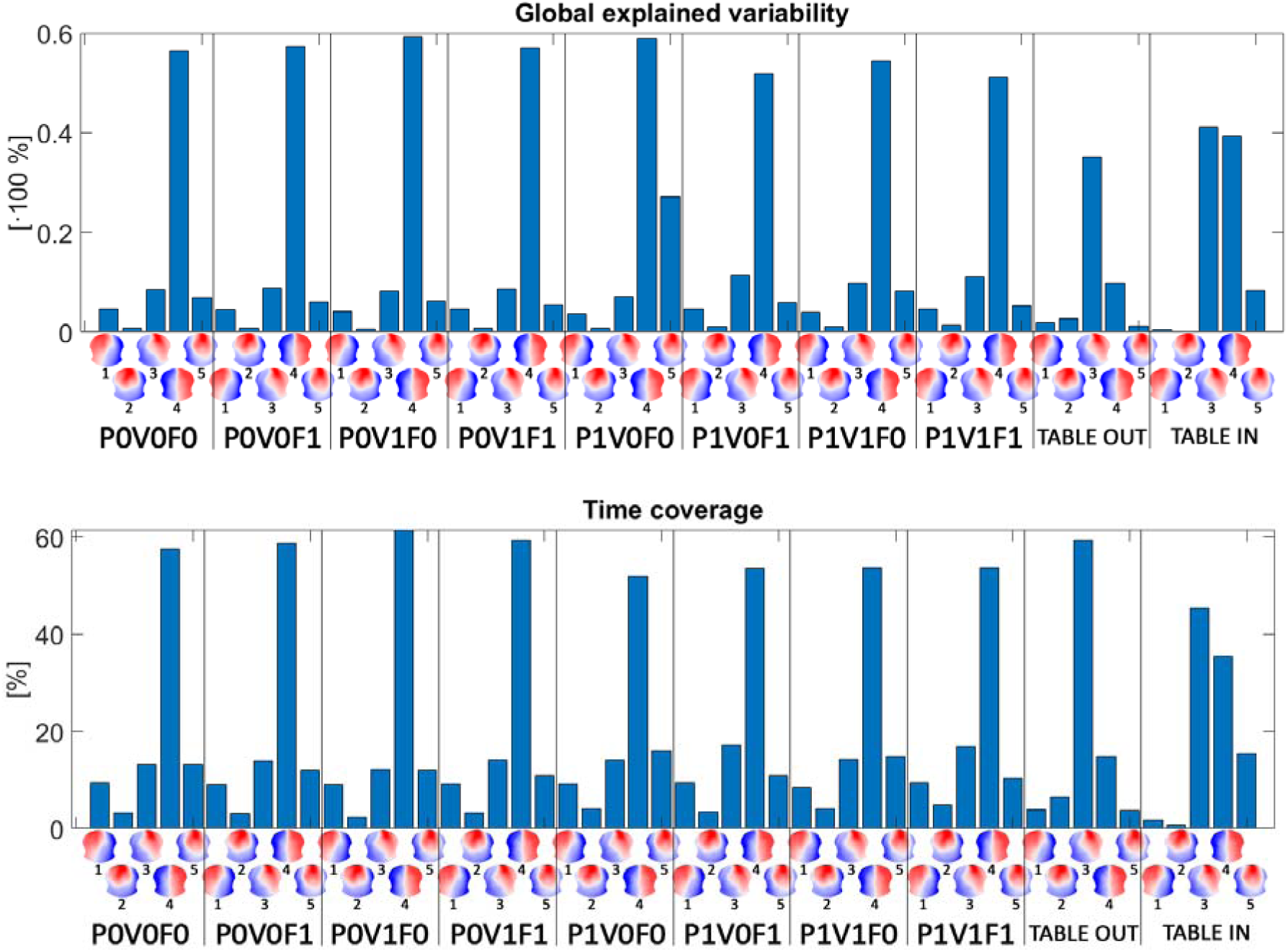
Comparison of each condition in the parameters of global explained variability (top) and time coverage (bottom). Each condition is labeled with codes: P = helium cooling pump, V = MR bore ventilation, F = fMRI protocol, 0 = OFF state and 1 = ON state. The last two conditions were recorded when the MR was in standby mode. VT (marked with 4) represents the sum of three similar VTs (PCM 4, 5, and 6 in Figure 3).

#### 3.2.3 Phantom dataset C

Analysis of PFL dataset revealed 8 stable states (Figure 5A) as optimum, which explained 99.84% of variability and 18.3% of signal was unlabeled, mainly in the last part of the measurement when the static magnetic field was already not present. Stable state PFL 7 explained most of the variability (59.1%) and covered 39.6% of signal. It shows spatial characteristics of VT. Spatiotemporal characteristics are listed in Suppl. table 8.

**Figure 5:**
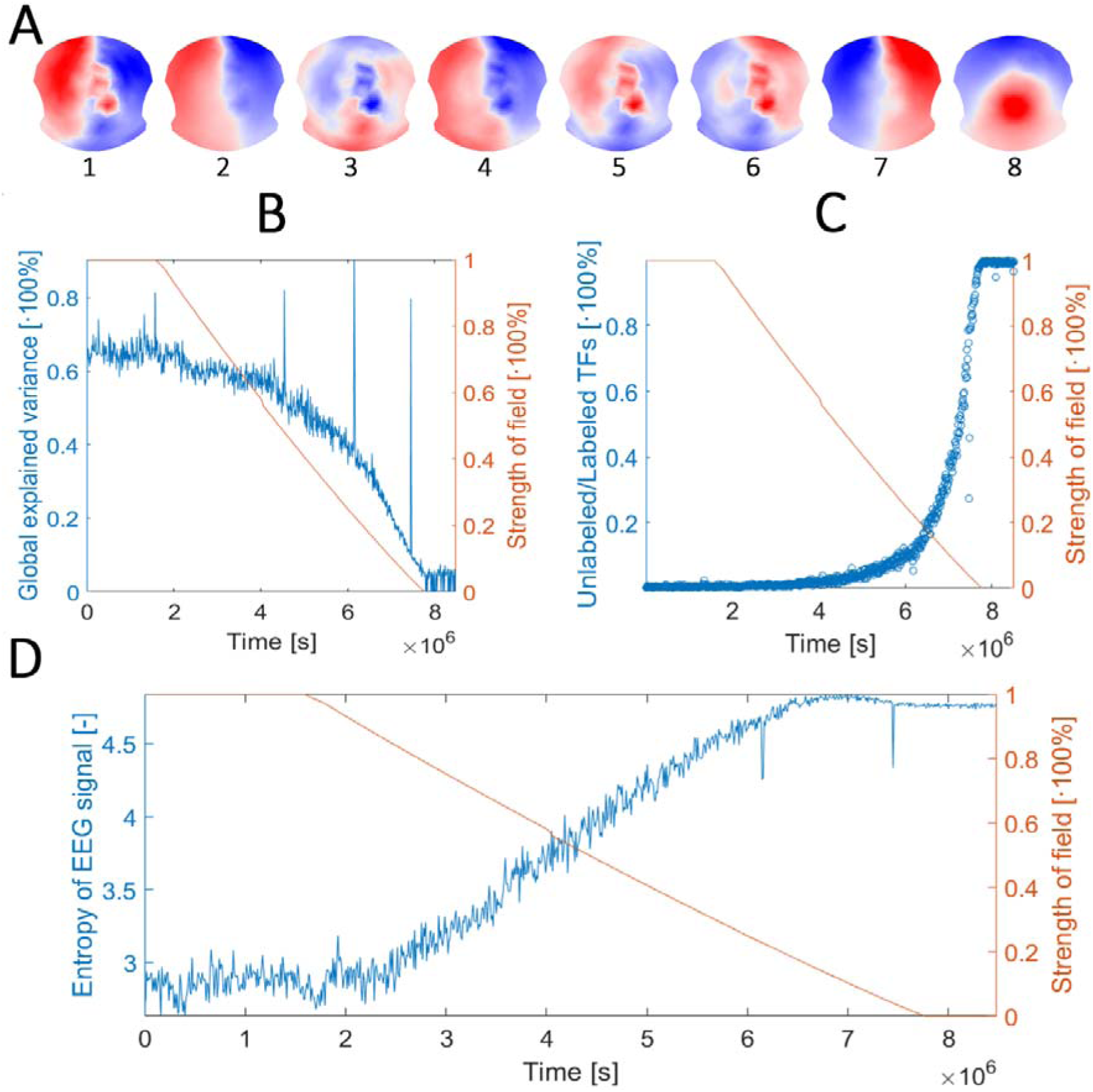
**A -** Resulting stable topographies for dataset PFL – phantom field lowering; **B –** Ratio between unlabeled and labeled TFs during lowering of the static magnetic field; **C –** Global explained variance of VT during lowering of the static magnetic field; **D –** Entropy of EEG signal during lowering of the static magnetic field

GEV of PFL 7 decreases with decreasing static magnetic field. As soon as the static field started to decrease, the GEV started to slowly decrease too. The GEV decrease accelerated when the static field lowered approx. to 50%. With no static magnetic field present, GEV of PFL 7 is below 10%. There are even windows with GEV of 0% in which there were no TF assigned to PFL 7. Events, where a technician physically interacted with MR machine are reflected by high peaks in GEV (Figure 5B).

Ratio between unlabeled and labeled TFs (Figure 5C) follows almost opposite course to a GEV.It is low as almost every TF is assigned to one of the representative stable topographies while the static field is high and starts to increase when the static field decrease below 20% and approaches 100% when static magnetic field was not present, as almost no TF is labeled with one of the representative stable topographies, i.e. there is no spatial structure in the data.

Entropy of signal (Figure 5D) is low in the beginning of the signal, where the static magnetic field was not lowering yet. When the static magnetic field begins to lower, entropy of EEG signal started to grow after several minutes and reaching its maximum when the static magnetic field is close to zero. During measurement of EEG without static magnetic field present, entropy remains high.

## 4. Discussion

The analysis of EEG microstates is a popular technique for exploring scalp EEG data with a focus on the behavior of large-scale brain networks and brain dynamics (Khanna et al., 2015; Michel and Koenig, 2018). The spatiotemporal parameters of the main prototypical microstate maps are evaluated in healthy subjects (Bréchet et al., 2019; Jabès et al., 2021; Liu et al., 2020), and their alterations are explored in the context of psychiatric (Nishida et al., 2013; Sverak et al., 2021) and neurological diseases (Lamoš et al., 2023, 2021). Some EEG/fMRI studies (Agrawal et al., 2022; Rajkumar et al., 2021; Schwab et al., 2015; Zappasodi et al., 2019) reported and assigned physiological significance to a topography with a distinct vertical line; we refer to this vertical topography as VT. Our study demonstrates that VT is partially a reflection of artifacts very likely caused by the movement. We support this claim with results from a series of EEG measurements performed outside and inside the MR environment with EEG net mounted either on human head or MR spherical phantom.

HSC EEG data were clustered into six microstates, four of them are very similar to the canonical microstates reported in the literature (HSC 1, 2, 5, and 6 to C (96.8%), A (98.5%), D (96.8%), and B (94.5%) respectively, correlation computed with Microstate template explorer (Koenig et al., 2024)). We found VT in HSC data in more than a half of subjects (52%, Suppl. figures 6, 7) although it was significantly less consistent when compared to HMR (Suppl. figure 10). On the other hand, we were able to identify epochs of VT with zig-zag character of the vertical line in continuous EEG signal (Suppl. figures 11, 12) which is at least a hint that some portion of VT may reflect artifacts also in HSC data. An identification of the VT’s source in HSC data was beyond the scope of this manuscript.

An analysis of HMR EEG data revealed six microstates. Three were similar to the canonical microstates (HMR 4, HMR 5, and HMR 6 to A (92.6%), B (85.7%), and C (87.6%) respectively), with HMR 6 being the most dominant one, in agreement with other microstate studies (Michel and Koenig, 2018; Zanesco, 2023). Our interest took HMR 1 with a strong vertical line in the middle, which covers almost 20% of the signal and explains 13.5% of GEV. The HMR 1 showed high prevalence (revealed in 92% of subjects, Suppl. figures 8, 9) and also high consistency at subject level (Suppl. figure 10). Moreover, we found that subjects who had higher FD (i.e. greater amount of movement during data acquisition), had also higher amount of VT in the EEG (Table 2, Figure 1). A clear VT can be also found directly in continuous EEG record during the movement of subject (Suppl. figure 15). In some EEG/fMRI studies, four clusters seems to be enough to reveal VT (Agrawal et al., 2022; Rajkumar et al., 2021). This was also case of our HMR EEG data (Suppl. figure 3 and Suppl. table 4) and it supports the hypothesis that the MR environment seems to increase the amount of VT in the data.

We also found that the shape of VT has some relationship to how the wires are placed and coupled on the EEG cap. A spatial filter applied during microstate analysis causes a straight vertical line; the same microstate revealed from un-filtered data shows that the vertical line has a “zigzag” character (Figure 6, Suppl. figure 17). This zig-zag character of vertical line can be also found directly in the continuous EEG data (Suppl. figure 15) as well as during the ICA (Suppl. figure 16). A closer look at the way the wires are routed either to the left or to the right side of the head confirms that it perfectly corresponds to the shape of the zigzag line. Commonly used EEG caps (mostly Brain Products, Munich, Germany or Electrical Geodesics Inc., Eugene, Oregon) have two main bundles of wires, one for each hemisphere (Figure 6, Suppl. figure 17). We suppose that this will be the case of every EEG cap which have bundles of wires separated on the left and right side. In the Suppl. material, we present several measurements with the BrainProducts 32-channel EasyCap to demonstrate, that the VT is not case of particular EEG cap. The fact, that VT seems not to be caused by a specific type of EEG cap or MR machine is also supported by literature, where authors identified VT as one of the dominant topographies. Suppl. table 9 shows that authors used various EEG caps and in case of simultaneous EEG/fMRI measurement also various MR machines.

**Figure 6:**
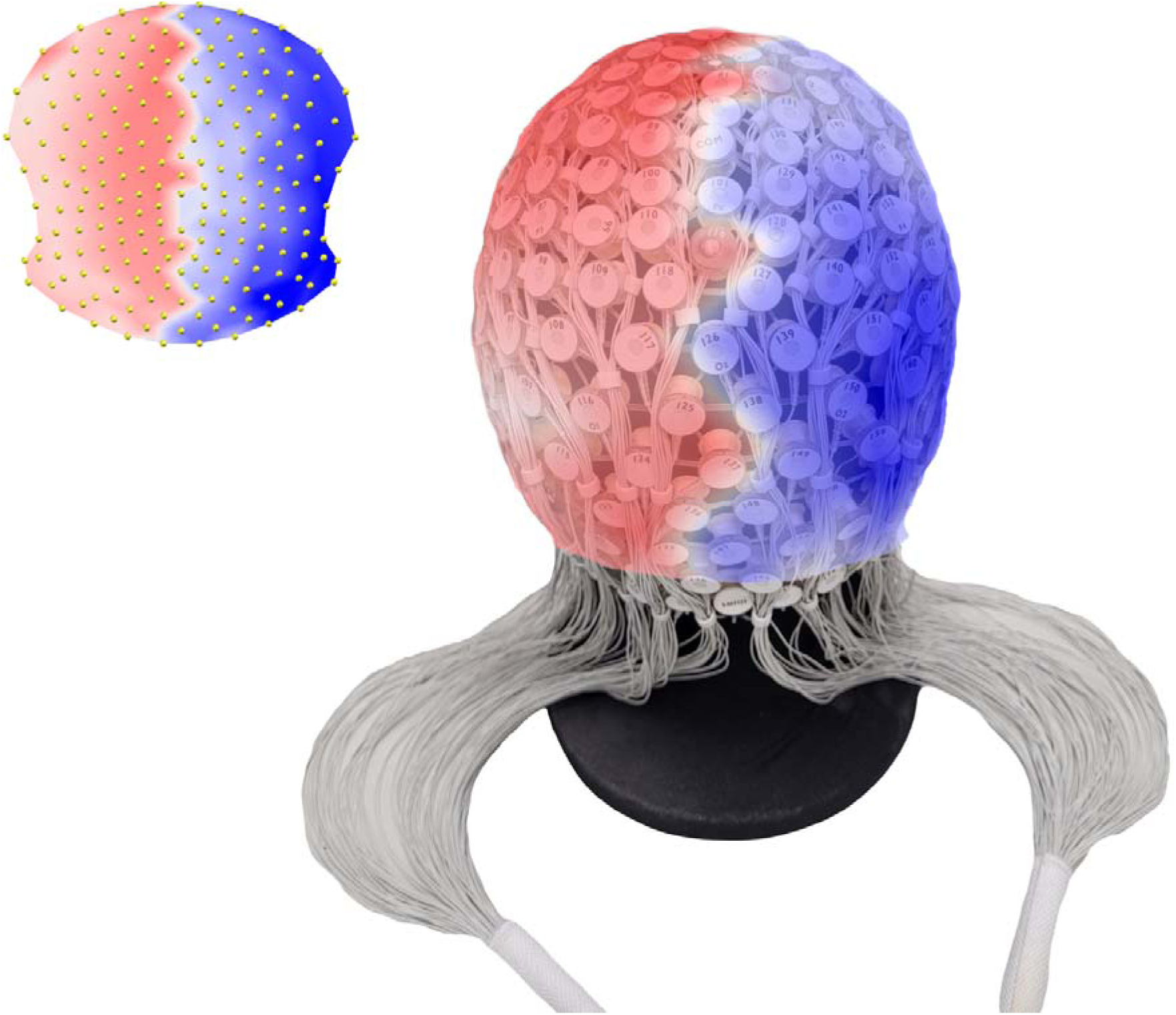
Spatially unfiltered VT is shown in the left corner of the figure. The map contains 204 electrodes, each of them symbolized by a yellow dot. On the right side of the figure is a real photograph of the 256-channel EGI EEG cap (mainly the occipital part) with VT mapped on the surface. It can be clearly seen that the vertical line follows the wiring and electrode placement.

An analysis of PMR EEG data showed that VT was the most dominant stable state and that several remaining topographies were contaminated with it. On the other hand, the PSC EEG data did not contain the VT at all. Taken together the above results, we think that the EEG data acquired in MR environment even after standard preprocessing steps are significantly polluted by an artifact that is reflected by the VT. The artifact results from the interaction of the magnetic field and the metal parts of the EEG system, very likely as a movement of the EEG cap within strong magnetic field.

We prepared another phantom measurement with ten different conditions to assess the influences of the helium cooling pump, MR bore ventilation, and MR gradient system on the VT presence in the data. The results show that neither MR gradients, nor the helium cooling pump, and MR bore ventilation have any effect on VT (Figure 4). The amount of VT decreased when the scanner was in standby mode and it further decreased when the phantom was outside the bore. Obviously, such scenario was close to the conditions in the shielded cabin, although there was still a low static magnetic field that could influence the data.

We also had a unique opportunity to investigate the effect of static magnetic field on the signal acquired by the EEG system during the process of reducing the static magnetic field to zero during the planned 3T MR Siemens Prisma machine upgrade. During the whole process, phantom EEG was recorded. As well as in other MR phantom measurements, VT is the most dominant topography. GEV of VT was decreasing during the lowering of the static magnetic field with accelerated decay as the field decreased below the half of its original strength. Also, the ratio between unlabeled and labeled TFs is minimal until the static magnetic field decreases to its 50%. Then it almost exponentially raises up to 100% when the static magnetic field disappears; this is very close to conditions in shielded cabin. Considering this, it seems VT remains stable until roughly 1.5 T magnetic field and thus we can hypothesize that VT is not only the case of 3T MR machine, but it will have similar properties in 3T and 1.5 MR machines. On the other hand, entropy of EEG signal starts to raise almost immediately with decreasing static magnetic field, thus the signal loses its spatial structure and becomes more random.

These two facts – that the amount of VT does not depend on the presence of known sources of artifacts and that it lowers when the MR is in standby mode and static magnetic field decreases – lead us to hypothesize that there is another unknown source of interaction between the static magnetic field and EEG system. It follows the Maxwell theory that the voltage magnitude induced in the wire loop in the magnetic field depends on the size and speed of change of either loop area or the magnetic field. The typical induced harmonic signal in our PCM data has amplitude of 10uV and frequency in the range of 0 to 50 Hz. If we consider two extreme cases, i.e. either perfectly constant static filed or completely motionless parts of EEG, such signal would be caused either by a harmonic change of loop area with the amplitude in the order of tenths of square millimeter in former case or by a harmonic change of the static filed in the order of units of micro-Tesla in the latter case (for details, please see Supplementary material). Such a change in the static magnetic field is far beyond manufacturer’s declarations. We thus hypothesize that the signal is caused by tiny vibrations either coming from MR machine or wider environment that causes tiny movements of EEG system in the strong static magnetic field.

Based on our results, we conclude that VT mainly reflects an artifact very probably caused by movement of various type. In shielded cabin the source of the movements would be the subject itself; e.g. movement of head and neck and blood pulsation may cause movement of sensors and thus slight changes in impedances of sensor-skin transition. In MR environment, the effect of movement is even magnified due to strong magnetic field and along with the known sources of movement (e.g. helium pump, air conditioning, subject), there are very probably other latent sources of vibrations of unclear origin.

At the same time, we cannot rule out, that some physiological activity may be reflected by topography similar to VT.

## 5. Conclusion

To conclude, we would like to highlight several points:

1) VT is present in PMR, PCM and PFL EEG data and it is the most dominant topography. Since the phantom definitely has no physiological activity, part of VT is certainly artificial.
2) The amount of VT is higher in HMR than in HSC. The significant increase is very likely related to the presence of the magnetic field. As an analysis of PFL data implies, amount of VT decreases with decreasing magnetic field.
3) Quantification and assessment of subject movement by FD confirms that VT was more dominant in EEG data if the participant moved more.
4) VT is present in PCM EEG data even when the fMRI protocol is not running. VT topography is not exclusively caused by fMRI sequence or by imperfection of AAS or by other known sources of artifacts. There has to be another source(s) of artifacts in the MR environment.
5) VT is not present in PSC EEG data; in cabin the phantom lies still on the chair without any movement or changes of magnetic field. The resulting topographies probably reflect environmental and device noise. The fact that 99% of signals were unlabeled implies that there is no pattern in the data. We were not able to identify any signs of VT even with the higher number of clusters.

We would like to emphasize that VT may significantly disrupt the results of the microstate analysis. As one of the resulting topographies, it is assigned to a portion of EEG samples and thus it necessarily biases the spatiotemporal characteristics of physiological microstates, the more so in the MR EEG data. These issues compromise follow-up analyses, such as microstate source localization or linking microstates to fMRI RSNs. In case of the shielded cabin EEG data, VT must be interpreted with caution after careful investigation of subject-level results. More investigation of VT will be needed to reduce its influence on the data. Our future directions will be aimed at finding a method that can lower the influence of VT so that microstate analysis is not affected by it. We will also investigate options of measuring protocols and measuring setup.

## Abbreviations

AAS: Average artifact subtraction
EEG: Electroencephalography
EEG/fMRI: Simultaneous electroencephalography and functional magnetic resonance measurement
FD: Framewise displacement
fMRI: Functional magnetic resonance imaging
GA: Gradient artifact
GEV: Global explained variance
GFP: Global field power
HD-EEG: High-density electroencephalography
HMR: Human magnetic resonance
EEG: dataset
HSC: Human shielded cabin
EEG: dataset
ICA: Independent component analysis
MR: Magnetic resonance
PA: Pulse artifact
PCM: Phantom magnetic resonance EEG dataset with ten different measuring conditions
PFL: Phantom magnetic resonance EEG dataset during lowering of static magnetic field
PMR: Phantom magnetic resonance EEG dataset
PSC: Phantom shielded cabin EEG dataset
RSN: Resting state network
VT: Vertical microstate topography

## Acknowledgements

We acknowledge the Multimodal and Functional Imaging Laboratory (MAFIL) core facility and the CEITEC MU institution, supported by the Czech-BioImaging large RI project (LM2023050 funded by MEYS CR), for their support with obtaining the scientific data presented in this paper. We would also like to thank to Mrs. Anne Johnson for correction of grammar and spelling.

## Compliance with ethical standards

Written informed consent was obtained from each participant before the study. The study was approved by the ethics committee of Masaryk University, Brno.

## Data availability

Due to institutional and ethical restrictions, the datasets used in the current study are only made available via a request to the corresponding author.

