## Supplementary_material for "Vertical topography in EEG microstates: Physiology or artifact manifestation?"

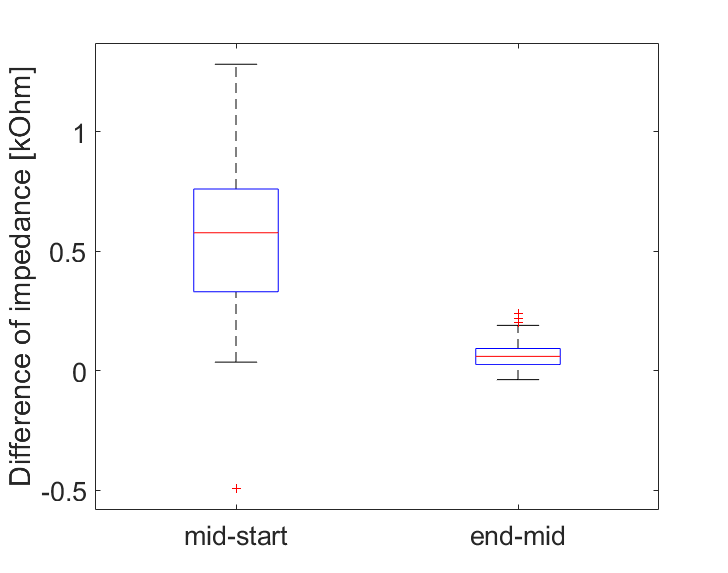

**Supplementary figure 1:** Impedances of electrodes during PFL EEG data recording were measured in three timepoints – in the beginning, middle and end of an experiment. The first boxplot shows differences between impedances measured in the middle and in the beginning of an experiment, the second boxplot shows differences between impedances measured in the end and in the middle of an experiment.

The fitting part of the microstate analysis produced four spatiotemporal parameters for each dataset: global explained variability, mean duration, time coverage, and occurrence. The tables for each dataset are listed below. Since 99% of the PSC EEG is unlabeled, quantification of its microstate spatiotemporal parameters is not reported here.

HSC dataset

**Supplementary table 1:** Microstate spatiotemporal parameters for human shielded cabin (HSC) EEG data, segmentation to 6 clusters

|  | GEV [%] | Mean duration [ms] | Time coverage [%] | Occurrence [counts/s] |
| --- | --- | --- | --- | --- |
| Map 1 | 24.23 ± 10.63 | 100.32 ± 18.34 | 30.74 ± 11.16 | 2.38 ± 0.47 |
| Map 2 | 8.10 ± 3.69 | 79.33 ± 7.31 | 14.82 ± 4.70 | 1.58 ± 0.37 |
| Map 3 | 4.19 ± 3.79 | 72.65 ± 10.45 | 11.52 ± 7.78 | 1.28 ± 0.72 |
| Map 4 | 3.14 ± 1.57 | 71.17 ± 4.57 | 7.97 ± 2.74 | 0.99 ± 0.31 |
| Map 5 | 13.93 ± 8.04 | 85.84 ± 12.65 | 21.00 ± 9.17 | 1.96 ± 0.63 |
| Map 6 | 7.75 ± 4.78 | 78.20 ± 6.00 | 13.96 ± 5.41 | 1.52 ± 0.49 |

**Supplementary table 2:** Microstate spatiotemporal parameters for human shielded cabin (HSC) EEG data, optimal segmentation to 4 clusters

|  | GEV [%] | Mean duration [ms] | Time coverage [%] | Occurrence [counts/s] |
| --- | --- | --- | --- | --- |
| Map 1 | 8.82 ± 4.51 | 78.17 ± 6.31 | 18.08 ± 5.64 | 1.92 ± 0.53 |
| Map 2 | 8.41 ± 3.31 | 78.21 ± 7.27 | 17.93 ± 4.91 | 1.92 ± 0.43 |
| Map 3 | 37.39 ± 11.06 | 122.22 ± 25.21 | 49.09 ± 10.55 | 2.99 ± 0.40 |
| Map 4 | 5.49 ± 4.73 | 72.00 ± 11.84 | 14.90 ± 9.79 | 1.63 ± 0.86 |

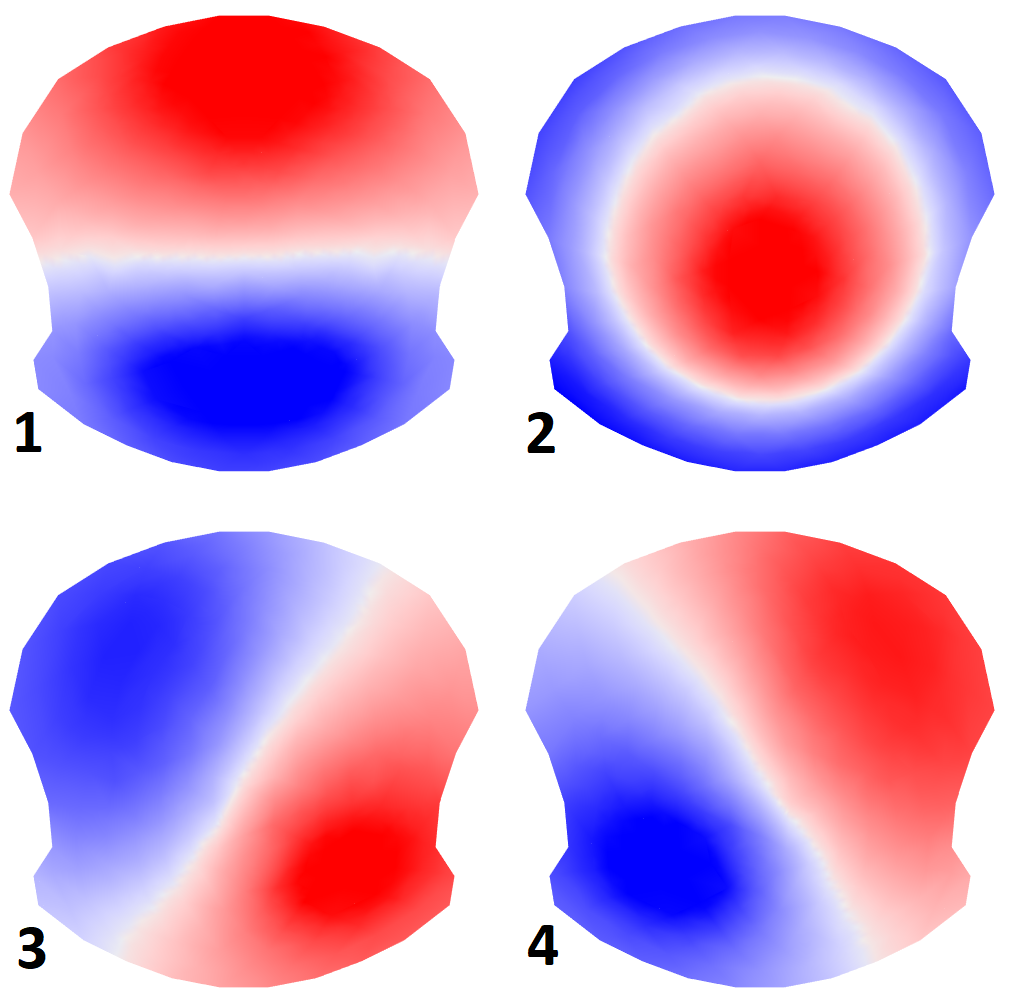

**Supplementary figure 2:** Resulting microstate topographies of human shielded cabin (HSC) EEG data – segmentation to 4 clusters

HMR dataset

**Supplementary table 3:** Microstate spatiotemporal parameters for human magnetic resonance (HMR) EEG/fMRI data, optimal segmentation to 6 clusters

|  | GEV [%] | Mean duration [ms] | Time coverage [%] | Occurrence [counts/s] |
| --- | --- | --- | --- | --- |
| Map 1 | 13.47 ± 11.71 | 78.31 ± 13.06 | 18.11 ± 11.39 | 1.89 ± 0.72 |
| Map 2 | 2.69 ± 2.24 | 65.57 ± 4.83 | 7.20 ± 4.39 | 0.96 ± 0.51 |
| Map 3 | 2.02 ± 1.47 | 64.51 ± 4.56 | 6.07 ± 3.24 | 0.83 ± 0.38 |
| Map 4 | 13.89 ± 6.43 | 79.15 ± 8.03 | 21.78 ± 6.88 | 2.36 ± 0.47 |
| Map 5 | 10.60 ± 4.91 | 75.19 ± 6.34 | 18.33 ± 6.31 | 2.10 ± 0.53 |
| Map 6 | 18.04 ± 9.22 | 87.90 ± 13.00 | 28.50 ± 11.06 | 2.63 ± 0.65 |

**Supplementary table 4:** Microstate spatiotemporal parameters for human magnetic resonance (HMR) EEG/fMRI data, segmentation to 4 clusters

|  | GEV [%] | Mean duration [ms] | Time coverage [%] | Occurrence [counts/s] |
| --- | --- | --- | --- | --- |
| Map 1 | 15.39 ± 12.01 | 78.70 ± 14.30 | 23.36 ± 11.92 | 2.44 ± 0.66 |
| Map 2 | 14.08 ± 6.57 | 77.17 ± 8.18 | 23.82 ± 7.46 | 2.63 ± 0.52 |
| Map 3 | 19.35 ± 9.69 | 86.69 ± 13.66 | 31.14 ± 12.19 | 2.90 ± 0.70 |
| Map 4 | 11.75 ± 4.86 | 74.24 ± 5.87 | 21.69 ± 6.18 | 2.52 ± 0.50 |

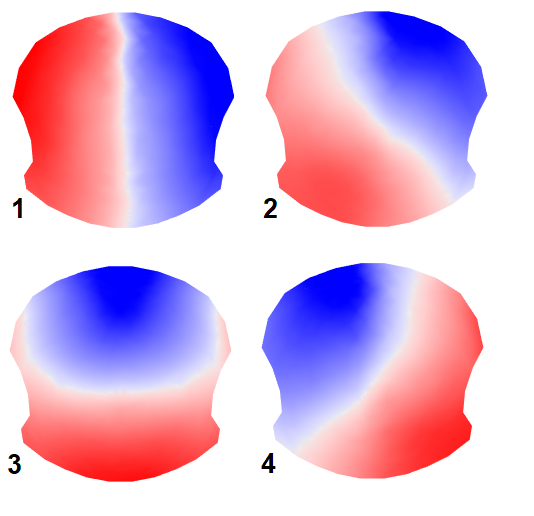

**Supplementary figure 3**: Resulting microstate topographies of human magnetic resonance (HMR) EEG/fMRI data – segmentation to 4 clusters

PMR dataset

**Supplementary table 5:** Stable state spatiotemporal parameters for phantom magnetic resonance (PMR) EEG/fMRI data recorded with same protocol as used for humans, segmentation to 6 clusters

|  | GEV [%] | Mean duration [ms] | Time coverage [%] | Occurrence [counts/s] |
| --- | --- | --- | --- | --- |
| Map 1 | 0,78 | 60,30 | 0,97 | 0,14 |
| Map 2 | 6,64 | 78,60 | 11,18 | 1,26 |
| Map 3 | 6,74 | 86,15 | 14,55 | 1,46 |
| Map 4 | 55,38 | 170,47 | 59,21 | 2,64 |
| Map 5 | 3,81 | 60,84 | 1,27 | 0,18 |
| Map 6 | 13,44 | 78,67 | 12,82 | 1,42 |

**Supplementary table 6:** Stable state spatiotemporal parameters for phantom magnetic resonance (PMR) EEG/fMRI data recorded with same protocol as used for humans, optimal segmentation to 8 clusters

|  | GEV [%] | Mean duration [ms] | Time coverage [%] | Occurrence [counts/s] |
| --- | --- | --- | --- | --- |
| Map 1 | 0.05 | 59.69 | 0.48 | 0.08 |
| Map 2 | 8.51 | 80.63 | 12.17 | 1.35 |
| Map 3 | 5.42 | 69.83 | 2.30 | 0.28 |
| Map 4 | 7.00 | 85.69 | 14.41 | 1.47 |
| Map 5 | 0.22 | 59.93 | 1.40 | 0.22 |
| Map 6 | 60.13 | 172.16 | 61.71 | 2.74 |
| Map 7 | 0.05 | 52.29 | 0.39 | 0.07 |
| Map 8 | 3.58 | 69.75 | 7.14 | 0.92 |

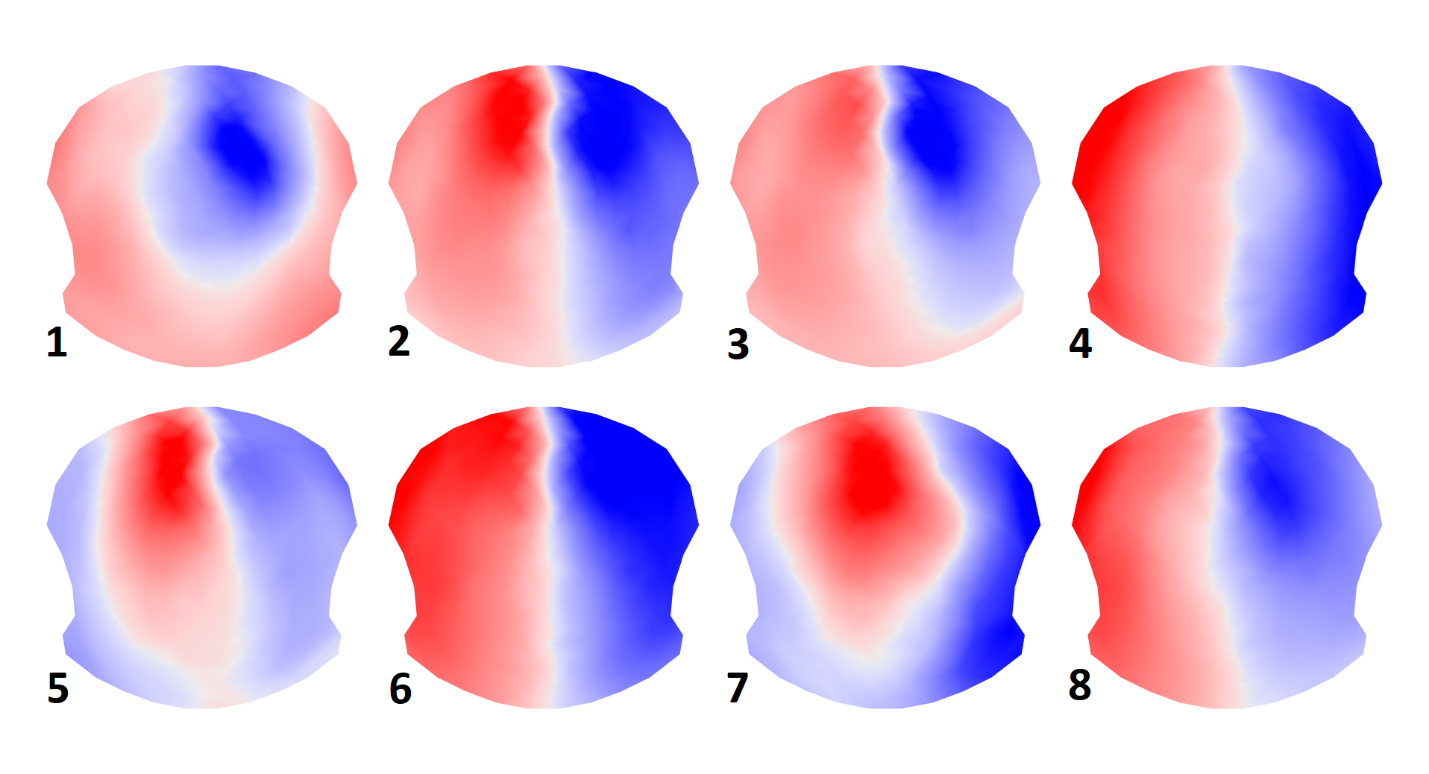

**Supplementary figure 4:** Resulting stable state topographies of phantom magnetic resonance (PMR) EEG/fMRI data recorded with same protocol as used for humans, optimal segmentation to 8 clusters

PSC dataset
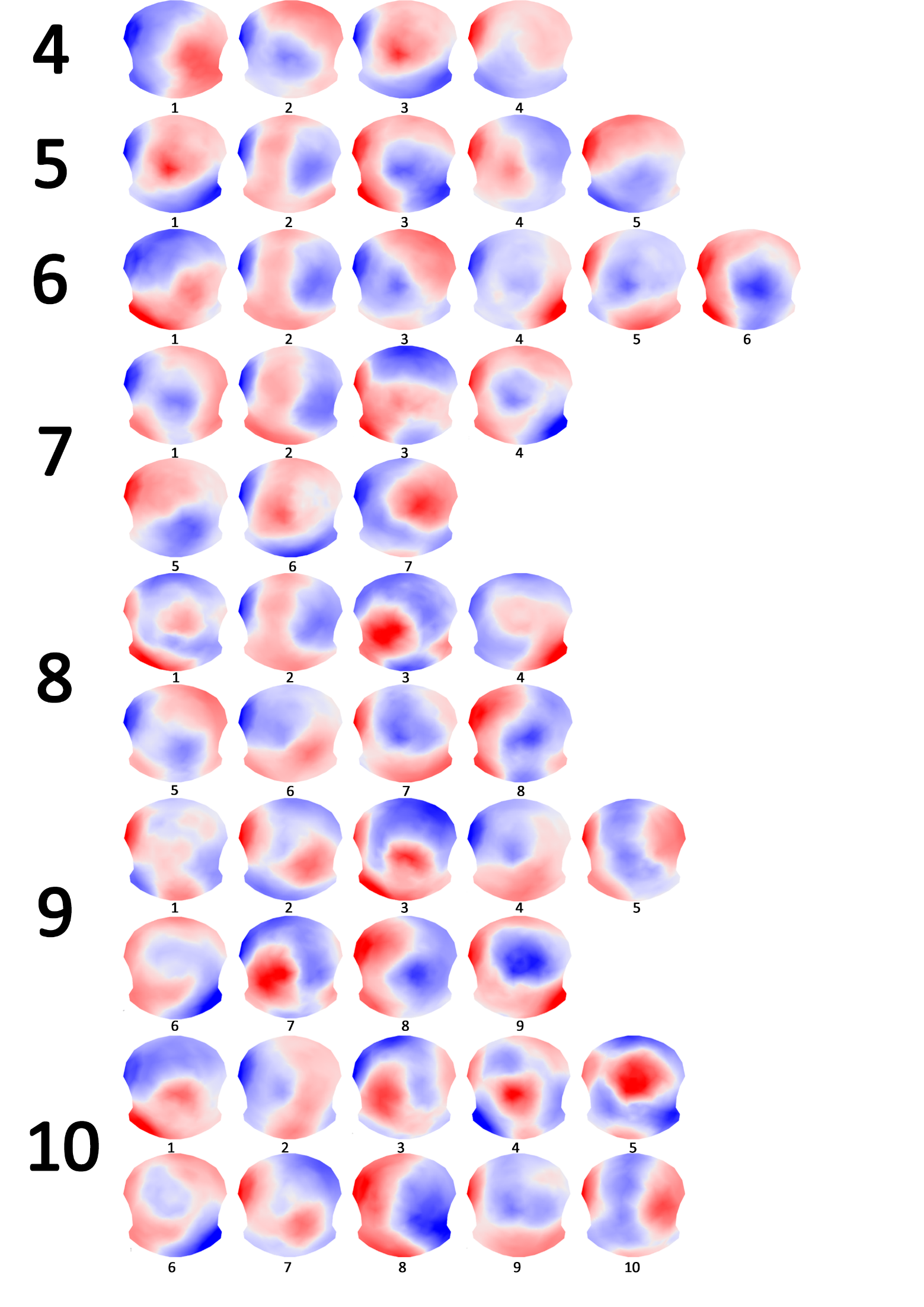
 **Supplementary figure 5:** Resulting stable topographies of phantom shielded cabin (PSC) EEG data, segmentations 4 to 10

PCM dataset

**Supplementary table 7:** Stable state spatiotemporal parameters for phantom conditions of measurement (PCM) EEG/fMRI data recorded with 10 different conditions

|  |  | Map 1 | Map 2 | Map 3 | Map 4 | Map 5 | Map 6 | Map 7 |
| --- | --- | --- | --- | --- | --- | --- | --- | --- |
| Helium pump OFF, ventilation OFF, fMRI OFF | GEV [%] | 4.70 | 0.70 | 8.51 | 12.79 | 6.90 | 36.76 | 1.17 |
|  | Mean duration [ms] | 77.72 | 67.85 | 83.46 | 86.35 | 80.26 | 118.02 | 64.20 |
|  | Time coverage [%] | 9.34 | 3.23 | 13.23 | 19.61 | 13.28 | 37.90 | 3.41 |
|  | Occurrence [N-times/s] | 1.07 | 0.44 | 1.40 | 2.00 | 1.48 | 2.66 | 0.48 |
| Helium pump OFF, ventilation OFF, fMRI ON | GEV [%] | 4.50 | 0.67 | 8.76 | 13.93 | 6.06 | 37.40 | 1.16 |
|  | Mean duration [ms] | 75.53 | 65.75 | 82.29 | 86.93 | 79.17 | 115.54 | 67.88 |
|  | Time coverage [%] | 9.12 | 3.04 | 13.80 | 20.93 | 11.97 | 37.78 | 3.36 |
|  | Occurrence [N-times/s] | 1.08 | 0.43 | 1.51 | 2.14 | 1.37 | 2.76 | 0.46 |
| Helium pump OFF, ventilation ON,  fMRI OFF | GEV [%] | 4.19 | 0.53 | 8.26 | 14.75 | 6.31 | 38.25 | 0.90 |
|  | Mean duration [ms] | 74.98 | 62.88 | 76.87 | 90.41 | 74.54 | 117.70 | 66.17 |
|  | Time coverage [%] | 8.99 | 2.46 | 12.27 | 22.03 | 12.01 | 39.38 | 2.87 |
|  | Occurrence [N-times/s] | 1.09 | 0.37 | 1.44 | 2.17 | 1.46 | 2.75 | 0.41 |
| Helium pump OFF, ventilation ON,  fMRI ON | GEV [%] | 4.60 | 0.68 | 8.66 | 13.56 | 5.49 | 37.87 | 1.60 |
|  | Mean duration [ms] | 75.93 | 65.33 | 82.09 | 86.24 | 77.38 | 117.68 | 67.73 |
|  | Time coverage [%] | 9.27 | 3.22 | 14.12 | 20.57 | 10.93 | 38.81 | 3.08 |
|  | Occurrence [N-times/s] | 1.12 | 0.46 | 1.54 | 2.10 | 1.27 | 2.77 | 0.42 |
| Helium pump ON, ventilation OFF,  fMRI OFF | GEV [%] | 3.67 | 0.73 | 7.13 | 9.13 | 27.08 | 22.81 | 1.50 |
|  | Mean duration [ms] | 77.52 | 67.43 | 83.83 | 89.40 | 82.41 | 110.43 | 70.70 |
|  | Time coverage [%] | 9.29 | 4.11 | 14.00 | 18.87 | 15.97 | 32.83 | 4.94 |
|  | Occurrence [N-times/s] | 1.08 | 0.57 | 1.49 | 1.87 | 1.72 | 2.49 | 0.65 |
| Helium pump ON, ventilation OFF,  fMRI ON | GEV [%] | 4.58 | 0.98 | 11.37 | 11.29 | 5.87 | 34.68 | 2.23 |
|  | Mean duration [ms] | 76.10 | 62.03 | 82.49 | 85.99 | 79.27 | 111.55 | 75.93 |
|  | Time coverage [%] | 9.42 | 3.48 | 17.14 | 17.35 | 10.83 | 36.21 | 5.57 |
|  | Occurrence [N-times/s] | 1.11 | 0.52 | 1.87 | 1.79 | 1.25 | 2.72 | 0.68 |
| Helium pump ON, ventilation ON,  fMRI OFF | GEV [%] | 3.92 | 1.01 | 9.82 | 12.56 | 8.24 | 33.64 | 1.82 |
|  | Mean duration [ms] | 75.71 | 71.27 | 81.36 | 84.11 | 84.00 | 112.41 | 67.39 |
|  | Time coverage [%] | 8.33 | 4.10 | 14.25 | 18.54 | 14.88 | 35.09 | 4.81 |
|  | Occurrence [N-times/s] | 0.99 | 0.53 | 1.56 | 1.96 | 1.59 | 2.64 | 0.66 |
| Helium pump ON, ventilation ON,  fMRI ON | GEV [%] | 4.62 | 1.38 | 11.16 | 11.51 | 5.34 | 34.36 | 1.96 |
|  | Mean duration [ms] | 77.37 | 73.21 | 84.56 | 85.53 | 79.91 | 115.97 | 72.53 |
|  | Time coverage [%] | 9.36 | 4.91 | 16.82 | 16.76 | 10.33 | 36.86 | 4.97 |
|  | Occurrence [N-times/s] | 1.10 | 0.60 | 1.75 | 1.76 | 1.16 | 2.65 | 0.63 |
| Standby mode, table outside | GEV [%] | 1.82 | 2.71 | 35.12 | 0.04 | 1.29 | 8.47 | 4.69 |
|  | Mean duration [ms] | 74.02 | 71.77 | 154.48 | 59.59 | 71.24 | 81.07 | 72.56 |
|  | Time coverage [%] | 3.93 | 6.53 | 59.34 | 0.23 | 3.70 | 14.63 | 11.65 |
|  | Occurrence [N-times/s] | 0.48 | 0.81 | 2.77 | 0.04 | 0.48 | 1.57 | 1.41 |
| Standby mode, table inside | GEV [%] | 0.41 | 0.08 | 41.15 | 0.97 | 8.33 | 30.01 | 0.28 |
|  | Mean duration [ms] | 61.93 | 50.42 | 119.29 | 66.27 | 76.73 | 102.76 | 56.70 |
|  | Time coverage [%] | 1.72 | 0.72 | 45.47 | 3.26 | 15.35 | 32.09 | 1.40 |
|  | Occurrence [N-times/s] | 0.26 | 0.14 | 3.07 | 0.46 | 1.81 | 2.58 | 0.23 |

**Supplementary table 8:** Stable state spatiotemporal parameters for phantom field lowering (PFL) EEG data

|  | GEV [%] | Mean duration [ms] | Time coverage [%] | Occurrence [counts/s] |
| --- | --- | --- | --- | --- |
| Map 1 | 5,68 ± 3,36 | 9,50 ± 0,44 | 24,40 ± 1,45 | 21,38 ± 0,10 |
| Map 2 | 22,71 ± 1,31 | 8,17 ± 0,23 | 15,45 ± 0,79 | 16,43 ± 1,55 |
| Map 3 | 0,21 ± 0,15 | 7,91 ± 0,35 | 2,87 ± 0,43 | 3,32 ± 0,31 |
| Map 4 | 2,61 ± 2,54 | 7,90 ± 0,41 | 6,31 ± 0,41 | 7,19 ± 0,93 |
| Map 5 | 0,68 ± 0,49 | 7,89 ± 0,37 | 5,65 ± 1,64 | 6,40 ± 1,49 |
| Map 6 | 0,44 ± 0,21 | 8,09 ± 0,42 | 5,47 ± 1,52 | 6,04 ± 1,27 |
| Map 7 | 59,12 ± 12,47 | 10,63 ± 0,23 | 39,59 ± 3,48 | 29,14 ± 2,79 |
| Map 8 | 0,77 ± 1,09 | 6,82 ± 1,87 | 0,27 ± 0,36 | 0,15 ± 0,18 |

Individual subject level topographies

The group level segmentation is done to obtain group specific topographies. Individual topographies of each subject contribute on the group topographies if they have spatially correlate (> 0.5) with the cluster. Below we present individual maps which contributed on the creation of the group map.

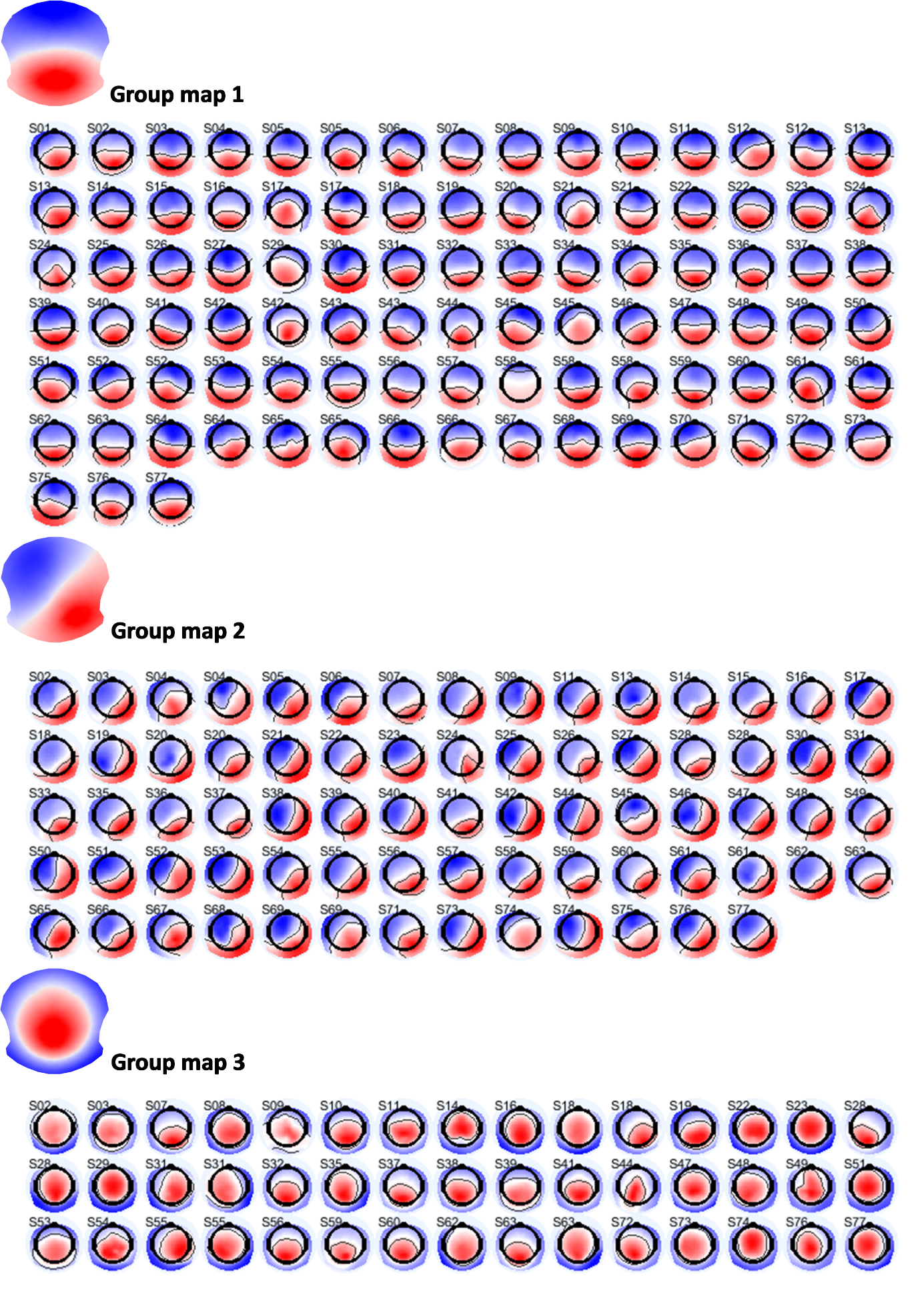

**Supplementary figure 6:** Individual subject topographies which contributed during the creation of the group topographies for the HSC dataset while clustering to the 6 clusters – part 1

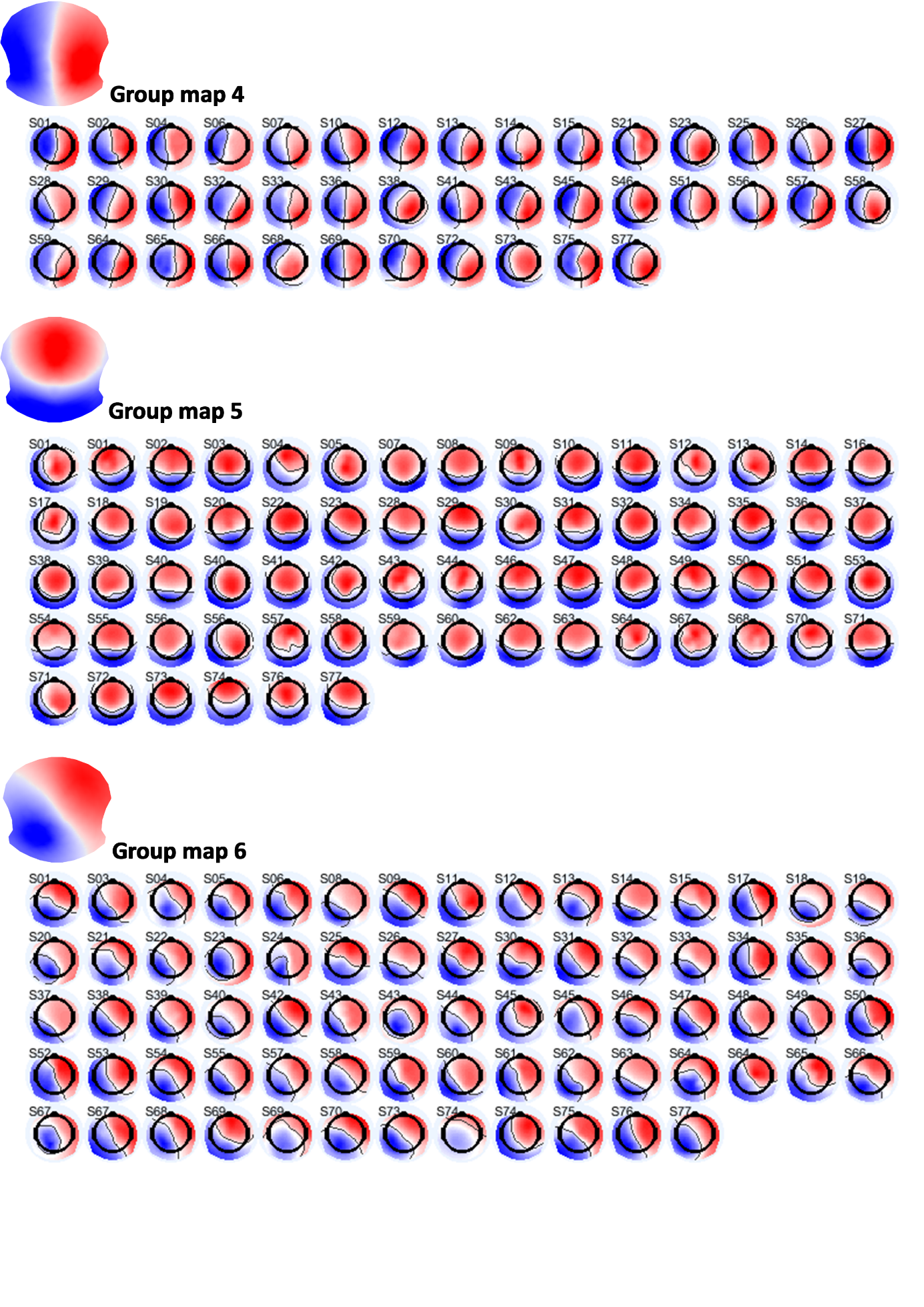

**Supplementary figure 7:** Individual subject topographies which contributed during the creation of the group topographies for the HSC dataset while clustering to the 6 clusters – part 2

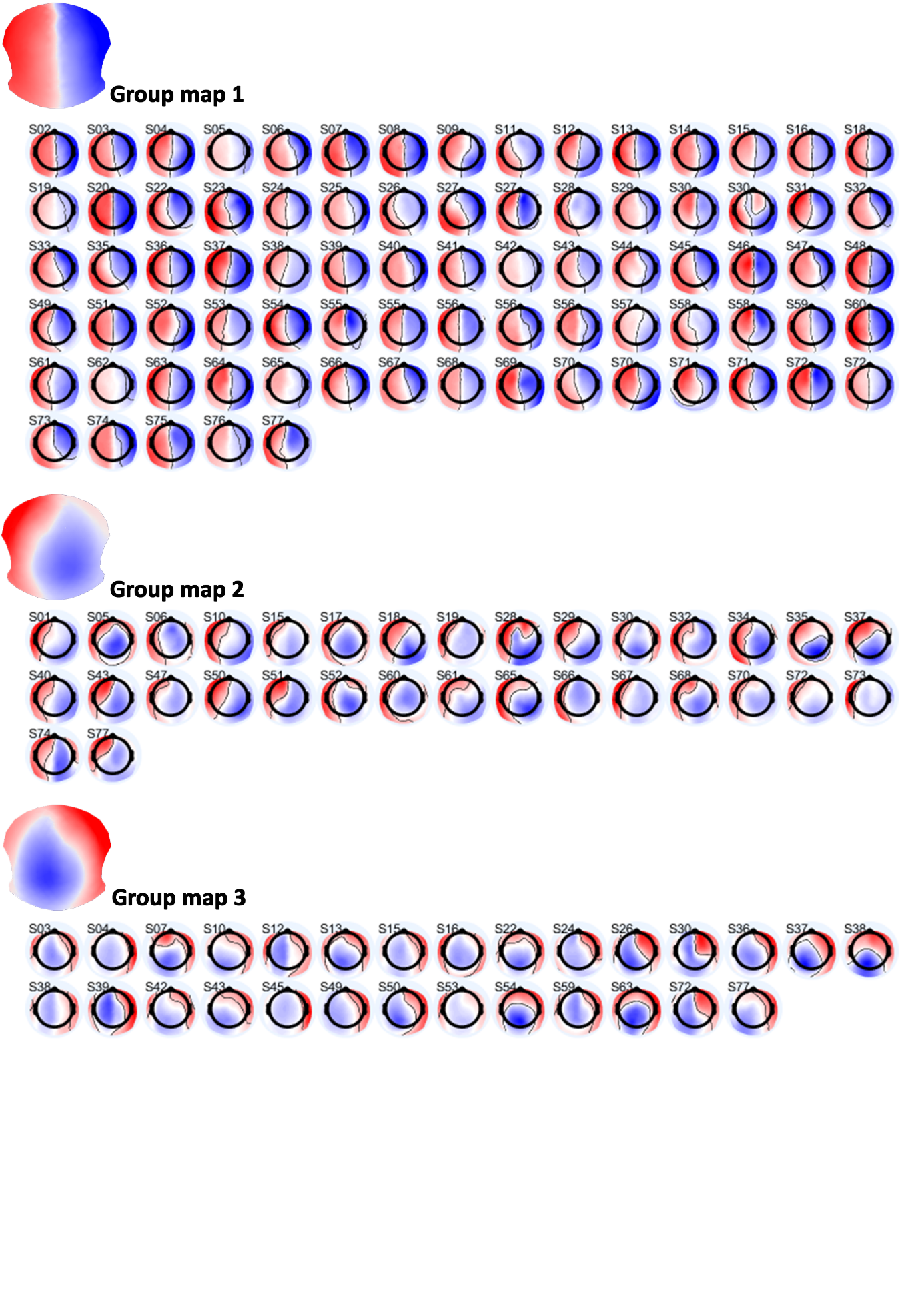
 **Supplementary figure 8:** Individual subject topographies which contributed during the creation of the group topographies for the HMR dataset while clustering to the 6 clusters – part 1

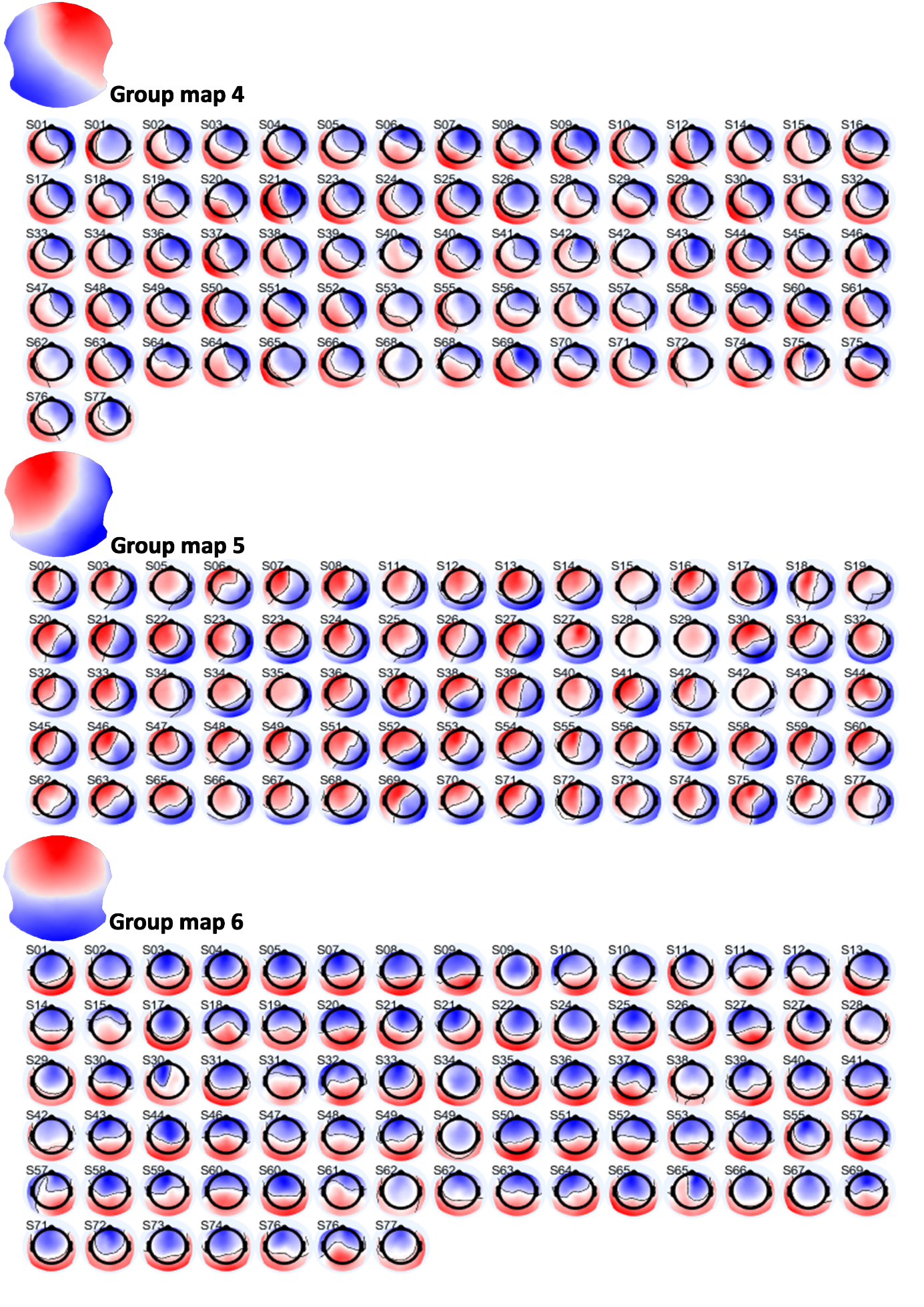

**Supplementary figure 9:** Individual subject topographies which contributed during the creation of the group topographies for the HMR dataset while clustering to the 6 clusters – part 2

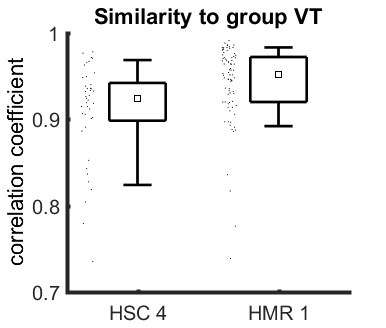

**Supplementary figure 10:** The similarity of subject-level VT to group-level VT is higher in HMR when compared to HSC data. The HSC 4 was revealed in 41 out of 77 subjects (53%). The HMR 1 was revealed in 71 out of 77 subjects (92%). The difference is significant at p<0.05 (p=0.0003, Man-Whitney test).

VT in continuous EEG signal

VT can be found in the continuous EEG signal, not only after the segmentation to the clusters. To demonstrate this, in the next four figures we show: 1) timeseries of selected channels, dominant group topographies are marked with corresponding color; 2) 2D representation of signal marked with the black vertical lines.

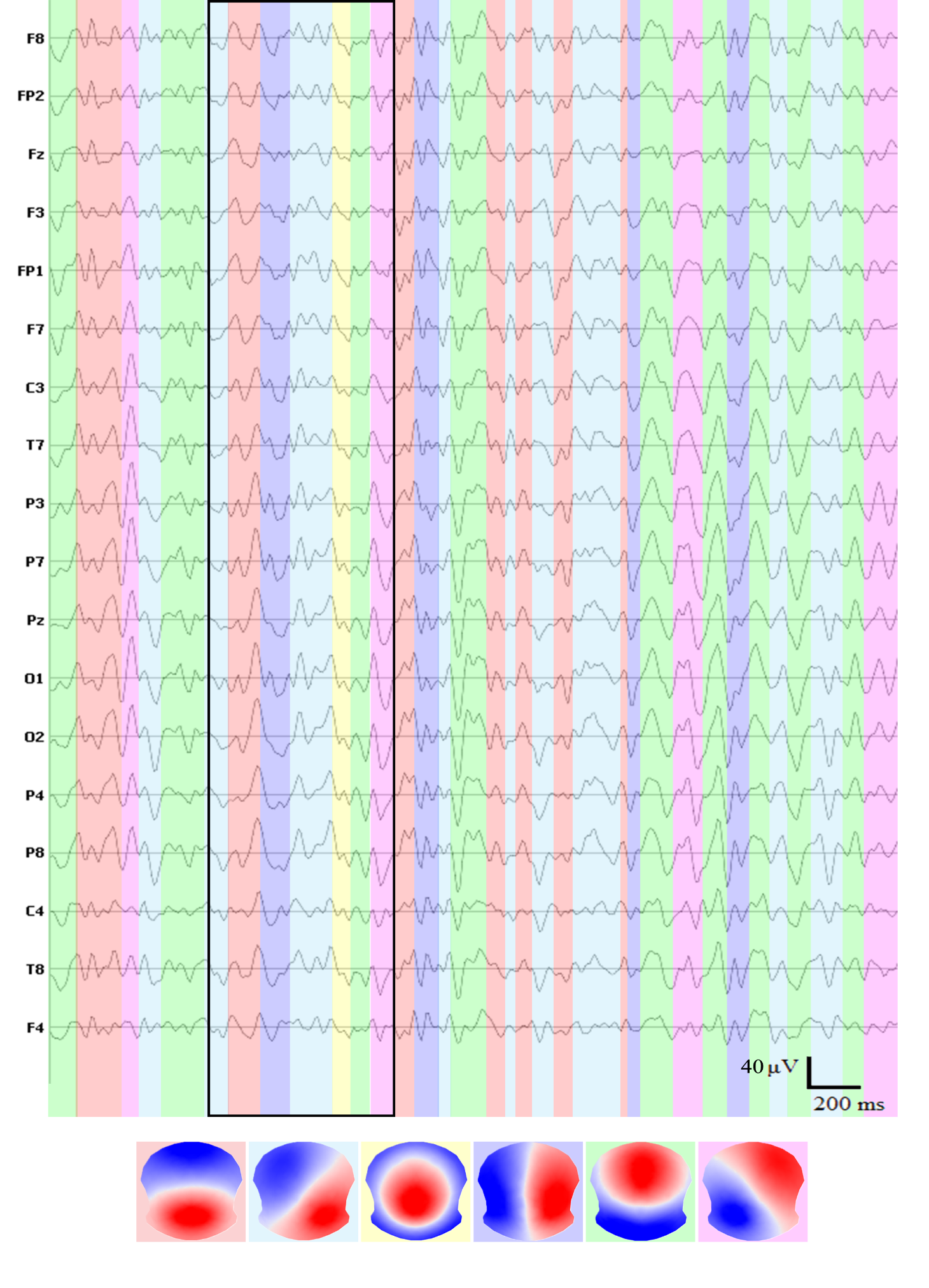

**Supplementary figure 11:** Example of continuous HSC EEG time series with fitted group topographies, subset of 18 channels. 2D projection of the signal marked with black lines is shown in the next figure – figure 8.

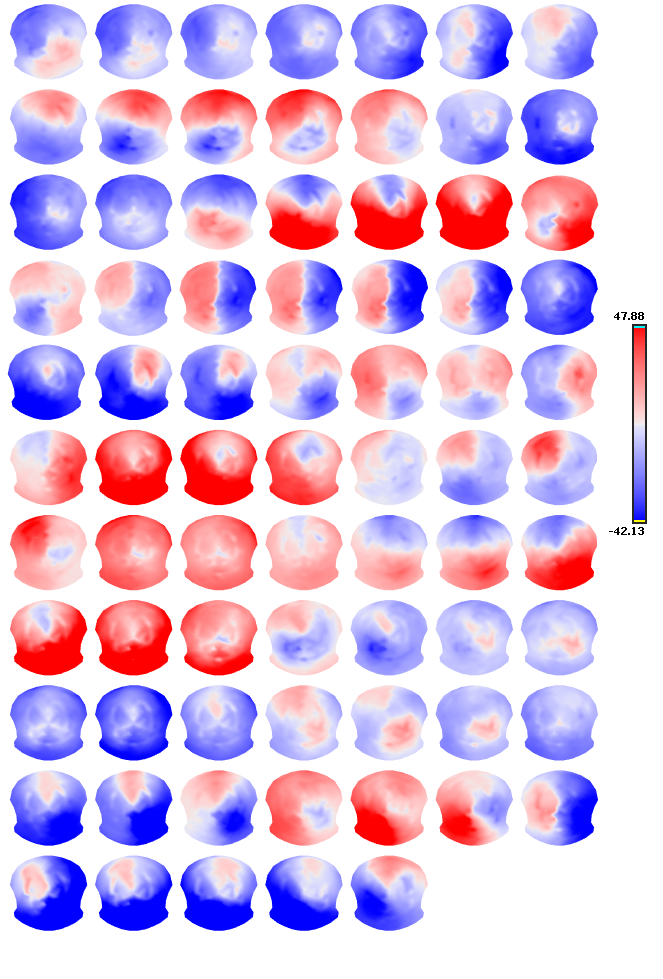

**Supplementary figure 12:** Example of the 2D representation of the HSC EEG data when VT is present (without spatial smoothing)

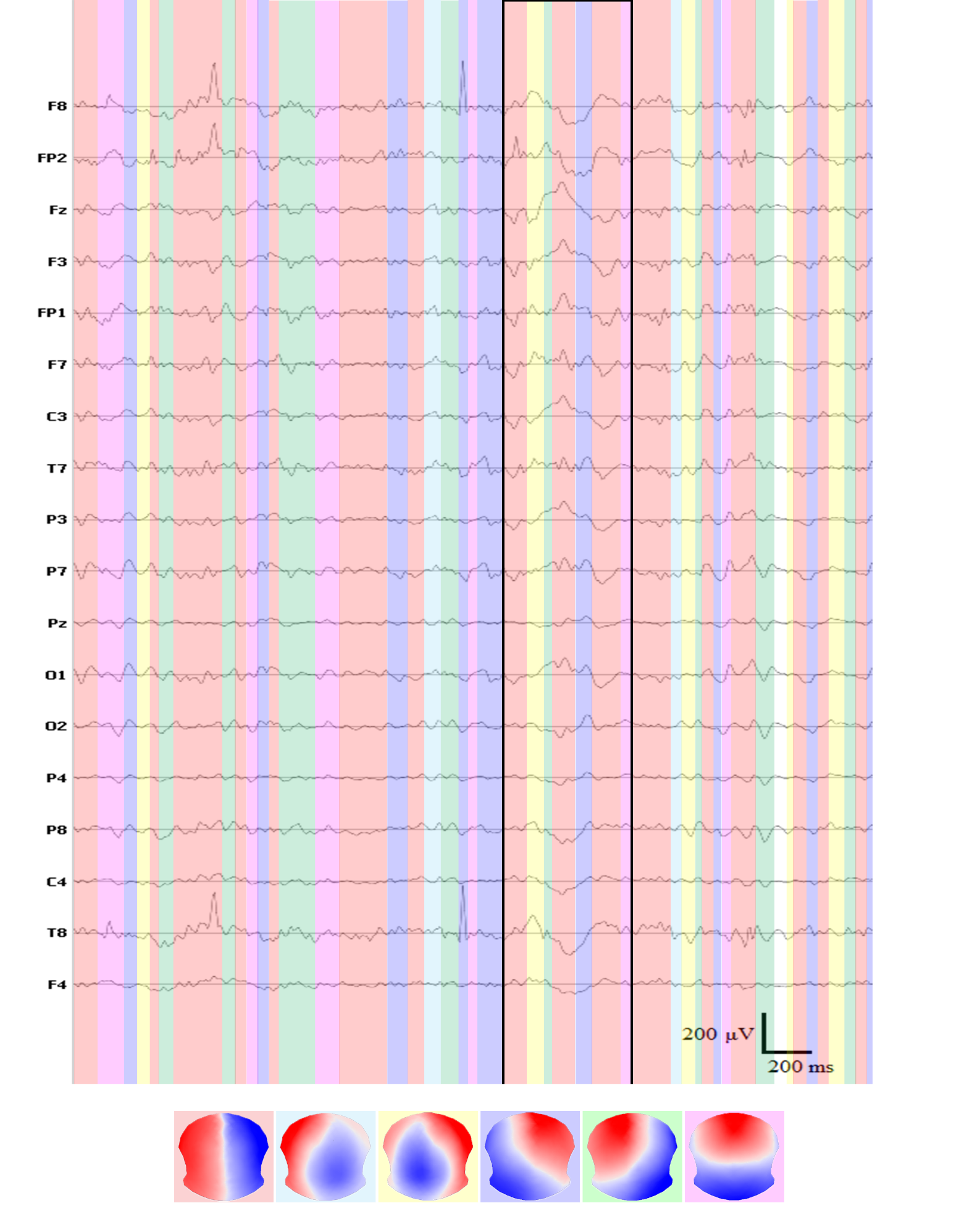
**Supplementary figure 13:** Example of continuous HMR EEG time series with fitted group topographies, subset of 18 channels. 2D projection of the signal marked with black lines is shown in the next figure – figure 6.

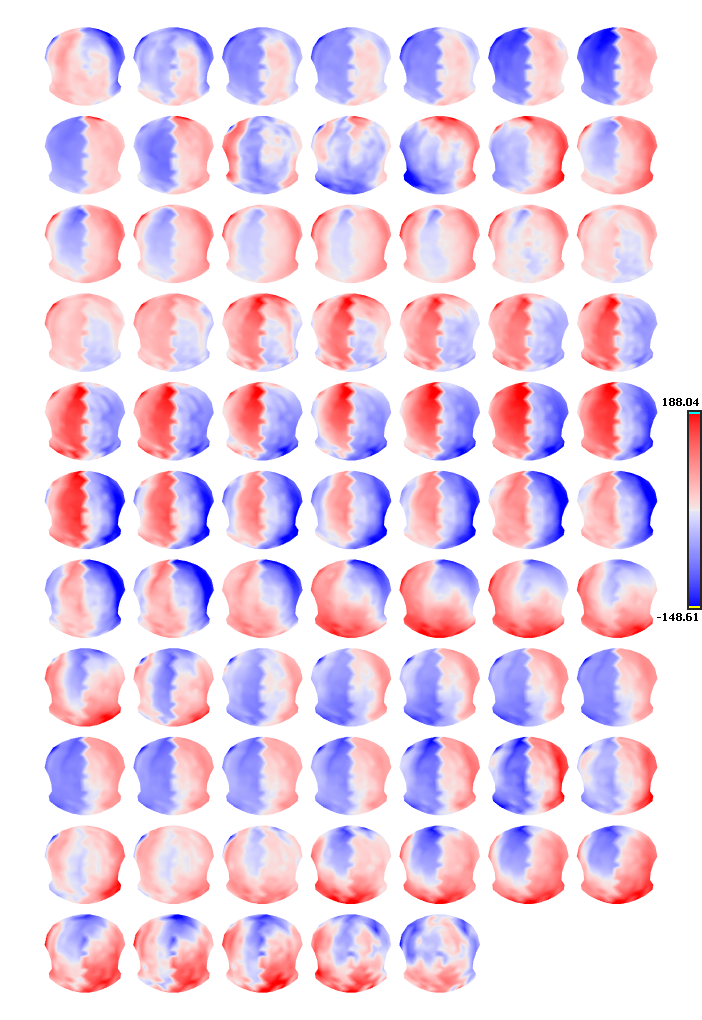

**Supplementary figure 14:** Example of the 2D representation of the HMR EEG data when VT is present (without spatial smoothing)

VT in continuous EEG signal during movement

VT can be identified in the continuous HMR EEG signal during movement of the subject as shown in the suppl. figure 9 below. Displayed topography is not spatially filtered, so the zig-zag character is evident.

| 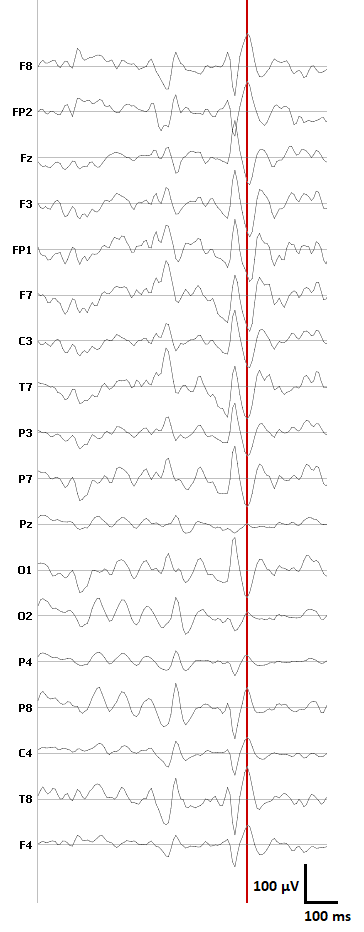 | 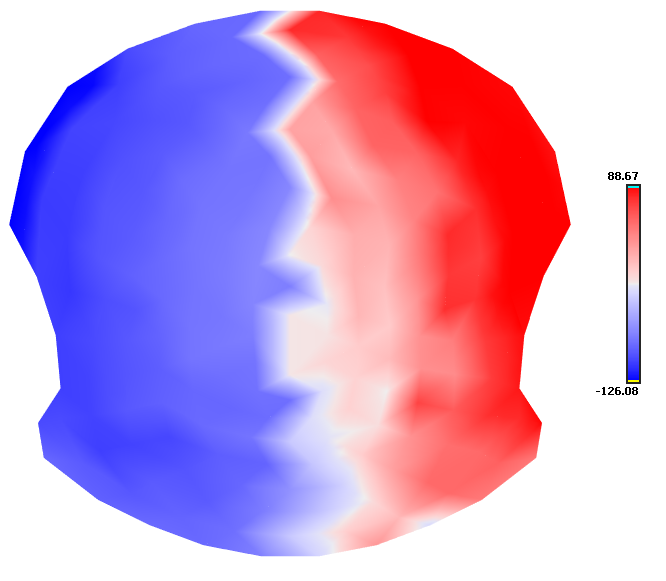 |
| --- | --- |

**Supplementary figure 15:** Example of the 2D representation of the HMR EEG data when VT is present during the movement of subject (without spatial smoothing).

In the introduction chapter, several articles present VT as one of the dominant topographies (on the group or subject level). Below, we present brief summary of the used recording equipment, preprocessing pipeline and microstate analysis setting in discussed papers. As it is evident, VT is not specific for the particular recording equipment or type of processing the data.

**Supplementary table 9:** Overview of the measuring devices, preprocessing and microstate pipelines used for acquiring and analyzing the simultaneous or classical EEG data in articles containing VT, which were mentioned in the introduction of the paper

| **Article** | **EEG cap** | **MR system** | **Preprocessing** | **MS analysis** | **Number of microstates** |
| --- | --- | --- | --- | --- | --- |
| **Agrawal et al., 2022** | BrainProducts 32-channel | Magnetom 3T, Siemens, TR 3000 ms | gradient and cardiobalistrogram correction, filtration 1–48 Hz, ICA, bad channels rejected, downsampling 250 Hz | GFP peaks, k-means | 5 frequency bands, 3 tasks, 4 MS each |
| **Bréchet et al., 2019** | BrainProducts 64-channel | - | filtration 1–40 Hz, ICA, interpolation, average reference, downsampling 250 Hz | GFP peaks, k-means individual + group level, temporal smoothing | 3 conditions, 6 MS each |
| **Custo et al., 2017** | EGI 256-channel | - | filtration 1–40 Hz, ICA, bad channel interpolation, average reference, downsampling 125 Hz | GFP peaks, k-means (100 runs individual level, 500 runs group level), temporal smoothing | 7 |
| **Rajkumar et al., 2021** | BrainProducts 32-channel | MR-BrainPET 3T, Siemens, TR 2250 ms | gradient and cardiobalistrogram correction, filtration 2–20 Hz, ICA, bad channels rejected, downsampling 1000 Hz | GFP peaks, AAHC individual + group level | 4 |
| **Schwab et al., 2015** | BrainProducts 96-channel | Magnetom Trio 3T, Siemens, TR 1980 ms | gradient and cardiobalistrogram correction, filtration 1–30 Hz, bad epochs rejected, downsampling 100 Hz | Topographic time frequency decomposition, k-means | 6 |
| **Thiele et al., 2023** | 28 active Ag/AgCl electrodes (10-20 system) | - | filtration 0.1–40 Hz, ICA, bad channel interpolation, average reference, downsampling 200 Hz | GFP peaks, k-means individual + group level | 5 |
| **Zappasodi et al., 2019** | Brainproducts 64-channel | Achieva 3T, Philips, TR 1950 ms | gradient and cardiobalistrogram correction, filtration 1–30 Hz, ICA, downsampling 125 Hz | GFP peaks, k-means individual + group level | 4 |
| **D’Croz-Baron et al., 2021** | EGI 65-channel | - | filtration 1–50 Hz, ICA, bad epochs rejection, average reference, downsampling 125 Hz | GFP peaks, k-means individual + group level | VT present on subject level, particular subject 4 MS |
| **Xu et al., 2020** | Brainproducts 64-channel | Prisma 3T, Siemens, TR 2000 ms | gradient correction, filtration 0.5–20 Hz, ICA, downsampling 250 Hz | GFP peaks, k-means individual + group level | VT present on subject level, 4 MS |

ICA decomposition – presence of VT

ICA decomposition often used for correction of the eye artifacts and ECG. In the figure below, first 30 independent components (ICs) obtained during the ICA decomposition of single subject’s EEG data are presented (timeseries and 2D representations). As is evident, several ICs are VT or are contaminated partially, implying that VT cannot be easily separated into one particular IC and thus easily corrected. Timeseries of VTs do not appear to be linked with known artefacts, so avoiding of all the VT ICs is speculative.

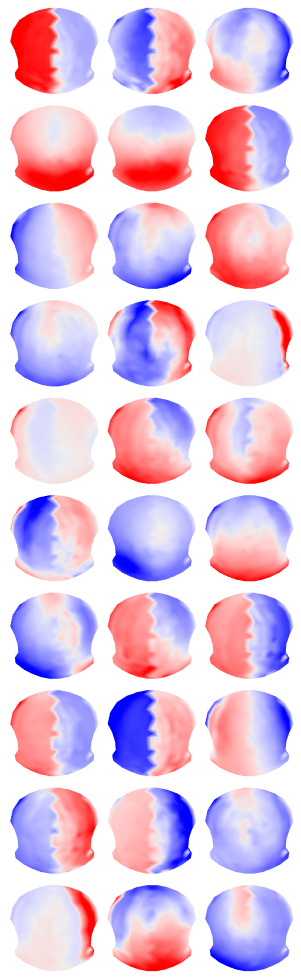

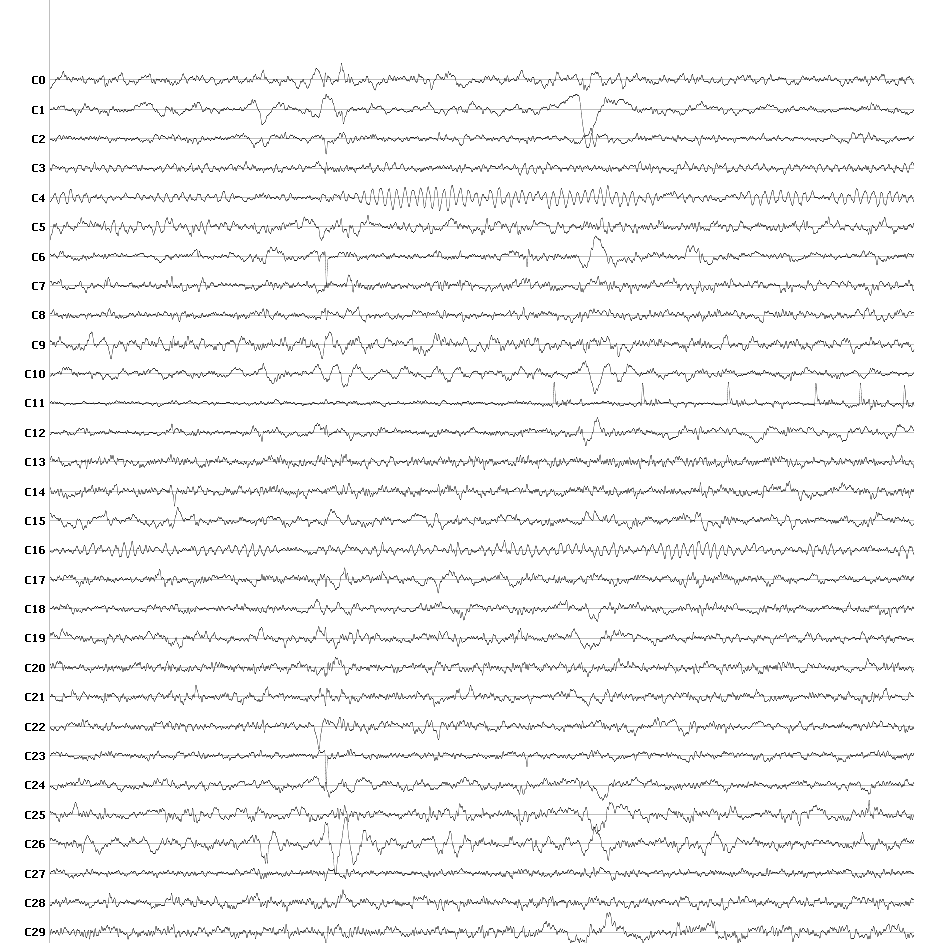

**Supplementary figure 16:** Timeseries and 2D representation of the first 30 independent components obtained during the preprocessing part of individual subject. It can be clearly seen, that several first topographies are VT and some others are contaminated with it.

Analysis of the data recorded with BrainProducts 32-channel EasyCap

Our analyses presented in the article are done on the data which were recorded with EGI 256-channel MR compatible cap. To avoid potential bias due to used EEG cap, we have measured phantom and human data with the 32-channel BP EasyCap in both – MR and shielded cabin – environment. The preprocessing of the data was the same as for the EGI cap, ECG and EOG channels were removed. One human and one phantom measurement was done just to illustrate that the presence of VT is not dependent on the specific type of EEG cap. Optimal number of microstates was selected based on the Metacriterion.

In human data, VT is present within four clusters in case of MR EEG data, and seven clusters in case of the cabin EEG data.

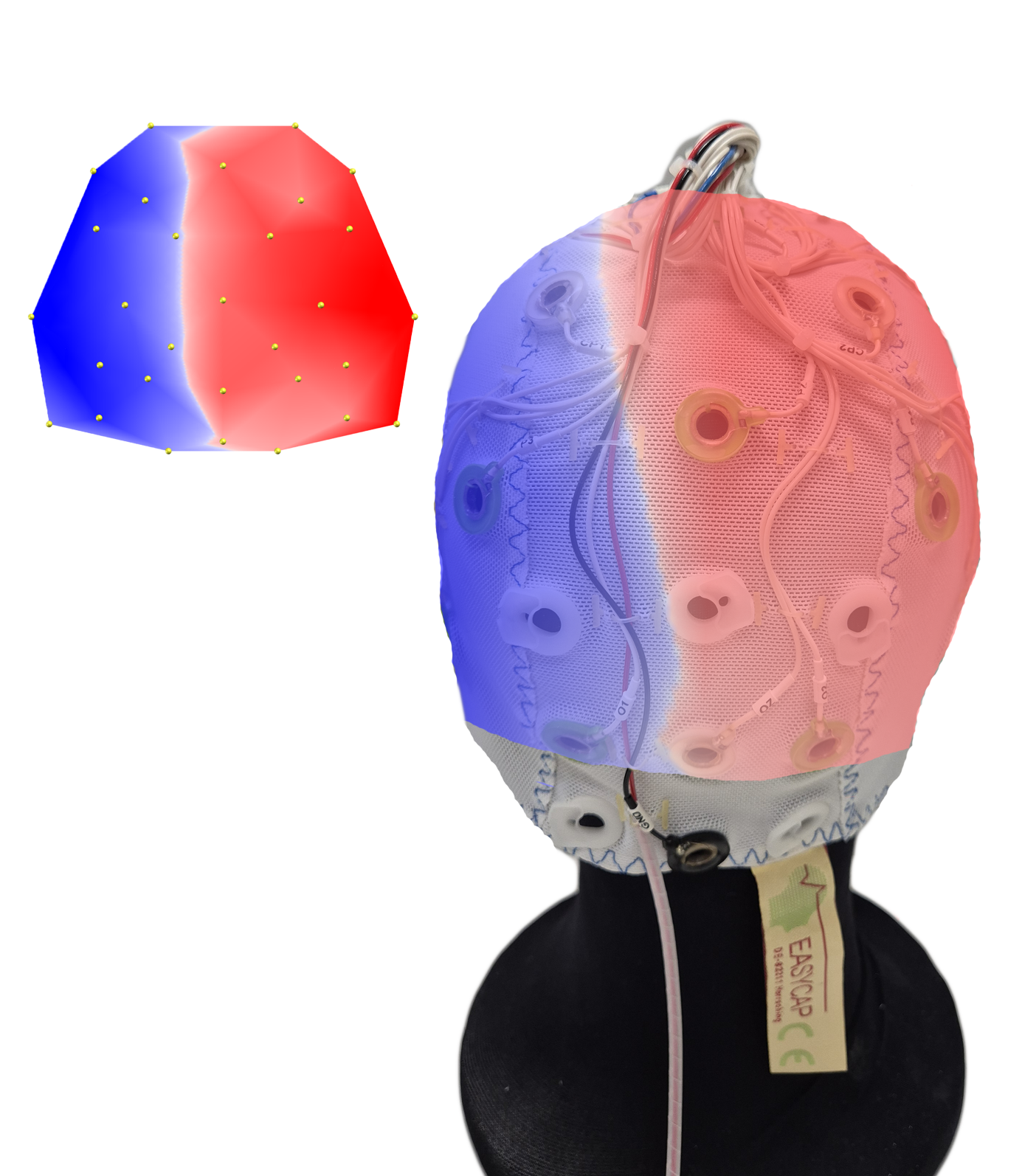

**Supplementary figure 17:** Spatially unfiltered VT is shown in the left corner of the figure. The map contains 30 electrodes, each of them symbolized by a yellow dot. On the right side of the figure is a real photo of the 32-channel BrainProducts Easy cap (mainly the occipital part) with VT mapped on the surface. It can be seen that the vertical line follows the electrode placement.

Human data

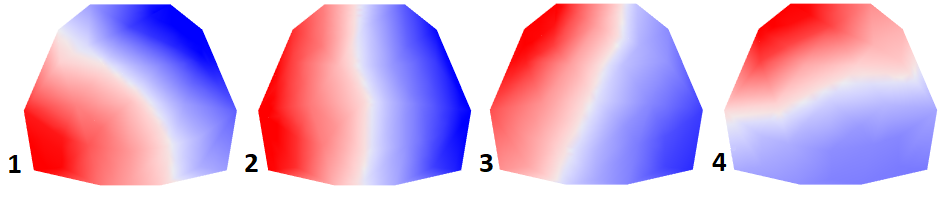

**Supplementary figure 18:** Resulting microstate topographies of human magnetic resonance EEG/fMRI recorded with 32-channel BrainProducts cap

**Supplementary table 10:** Microstate spatiotemporal parameters for human MR EEG data recorded with 32-channel BrainProducts cap

|  | GEV [%] | Mean duration [ms] | Time coverage [%] | Occurrence [counts/s] |
| --- | --- | --- | --- | --- |
| Map 1 | 11,46 | 73,13 | 18,95 | 2,27 |
| Map 2 | 30,12 | 100,32 | 36,99 | 2,97 |
| Map 3 | 17,66 | 82,44 | 25,47 | 2,67 |
| Map 4 | 25,02 | 81,74 | 18,58 | 1,82 |

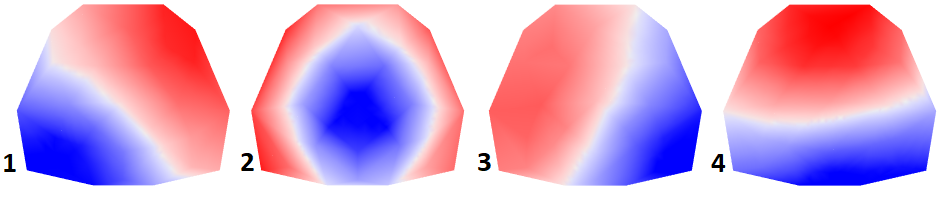

**Supplementary figure 19:** Resulting microstate topographies of human shielded cabin EEG data recorded with 32-channel BrainProducts cap

**Supplementary table 11:** Microstate spatiotemporal parameters for human shielded cabin EEG data recorded with 32-channel BrainProducts cap

|  | GEV [%] | Mean duration [ms] | Time coverage [%] | Occurrence [counts/s] |
| --- | --- | --- | --- | --- |
| Map 1 | 15,72 | 82,19 | 27,00 | 2,80 |
| Map 2 | 4,67 | 67,49 | 12,95 | 1,73 |
| Map 3 | 12,31 | 77,93 | 22,90 | 2,55 |
| Map 4 | 26,24 | 93,05 | 37,15 | 3,34 |

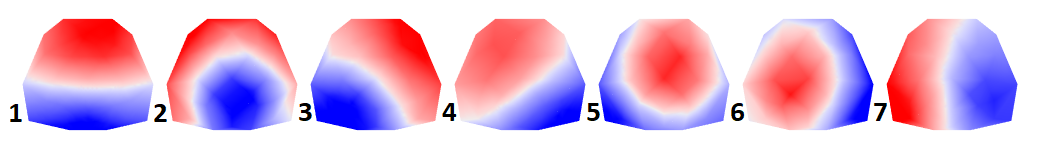

**Supplementary figure 20:** Resulting microstate topographies of human shielded cabin EEG data recorded with 32-channel BrainProducts cap – segmentation to 7 clusters

**Supplementary table 12:** Microstate spatiotemporal parameters for human shielded cabin EEG data recorded with 32-channel BrainProducts cap, segmentation to 7 clusters

|  | GEV [%] | Mean duration [ms] | Time coverage [%] | Occurrence [counts/s] |
| --- | --- | --- | --- | --- |
| Map 1 | 15,98 | 84,81 | 20,32 | 2,11 |
| Map 2 | 5,28 | 74,82 | 10,95 | 1,32 |
| Map 3 | 9,94 | 78,08 | 16,25 | 1,84 |
| Map 4 | 11,69 | 80,97 | 17,73 | 1,95 |
| Map 5 | 6,72 | 75,57 | 12,69 | 1,49 |
| Map 6 | 4,49 | 71,56 | 9,51 | 1,20 |
| Map 7 | 6,27 | 76,25 | 12,56 | 1,47 |

Phantom data

32-channel BrainProducts cap was mounted on a spherical MR phantom (same process as with the EGI cap) and put into the MR system, two conditions were recorded (approx. 5 minutes of recording) – during fMRI protocol running and without fMRI protocol running. Individual stable topographies were almost the same for both conditions, group level stable topographies are present below.

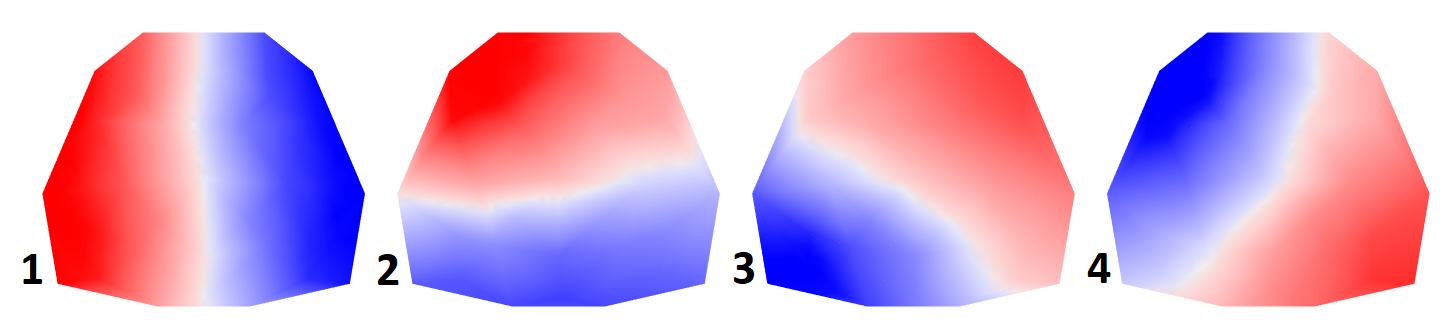

**Supplementary figure 21:** Resulting stable topographies of phantom magnetic resonance EEG/fMRI data recorded with 32-channel BrainProducts cap

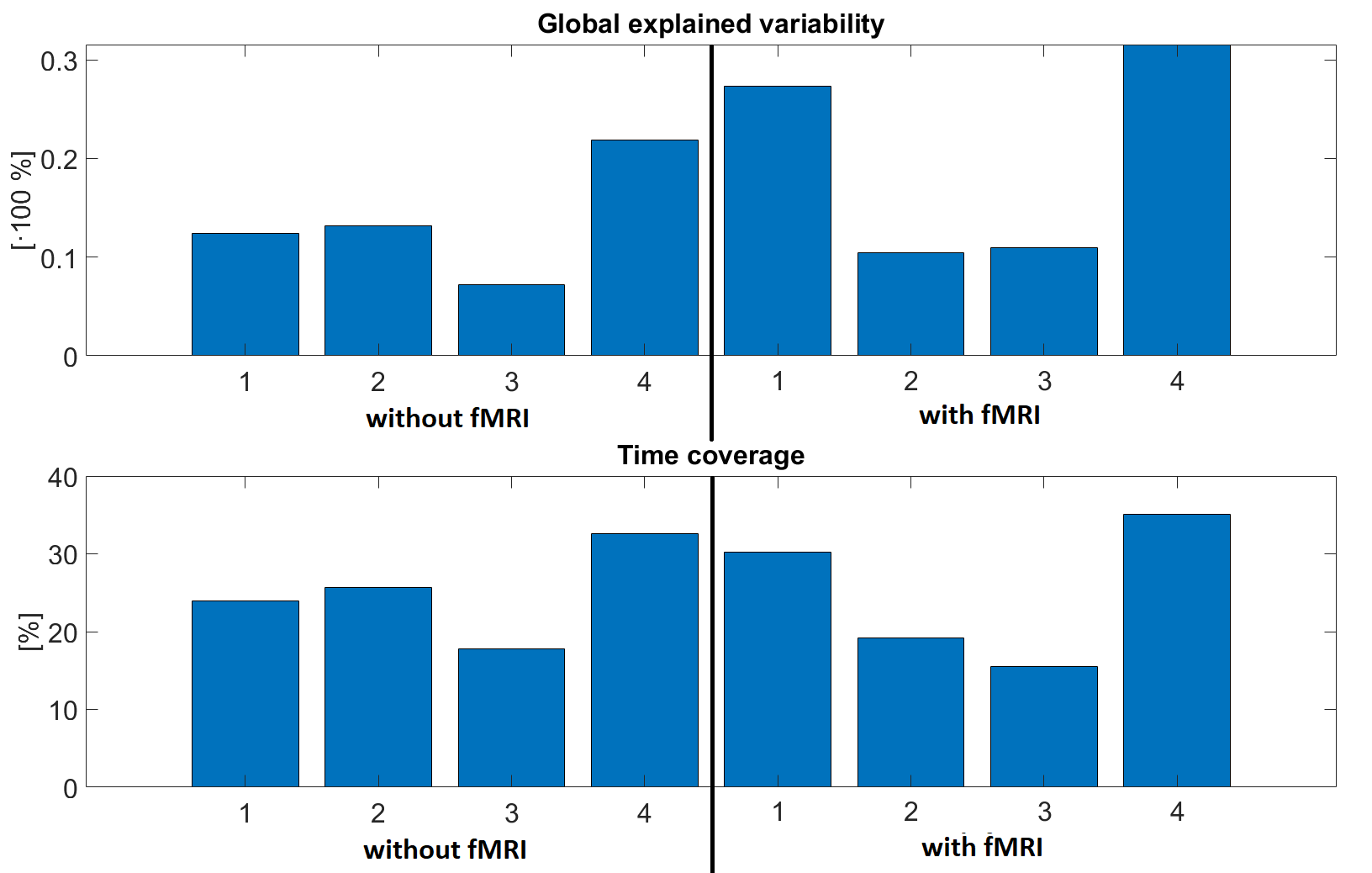

**Supplementary figure 22:** Spatiotemporal parameters of stable topographies of phantom magnetic resonance EEG/fMRI data recorded with 32-channel BrainProducts cap

Data from a shielded cabin were also recorded with the 32-channel BrainProducts cap, approx. 5 minutes of recording. When the spatial filter was used, 7 stable states were obtained and fitted back to the data. Since the 98 % of the data was labeled with one of the stable topographies, they do not represent the data well as the GEV for each of the stable states is really low.

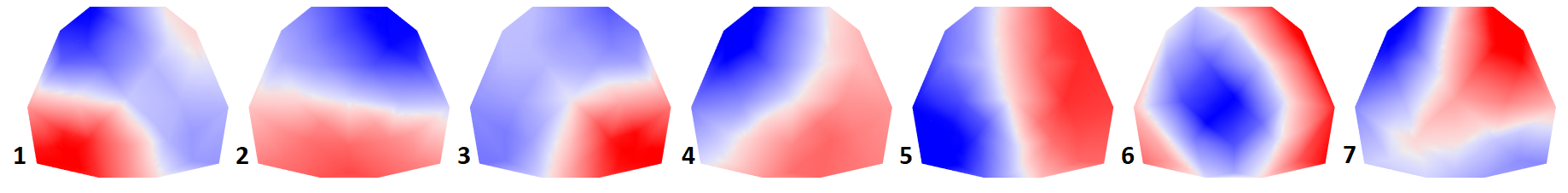

**Supplementary figure 23:** Resulting stable topographies of spatially filtered phantom shielded cabin EEG data recorded with 32-channel BrainProducts cap

**Supplementary table 13:** Stable state spatiotemporal parameters for phantom shielded cabin EEG data recorded with 32-channel BrainProducts cap

|  | GEV [%] | Mean duration [ms] | Time coverage [%] | Occurrence [counts/s] |
| --- | --- | --- | --- | --- |
| Map 1 | 5,30 | 90,92 | 17,78 | 1,73 |
| Map 2 | 3,37 | 85,10 | 12,28 | 1,30 |
| Map 3 | 6,10 | 94,18 | 20,01 | 1,86 |
| Map 4 | 5,77 | 94,36 | 17,95 | 1,65 |
| Map 5 | 2,08 | 83,34 | 9,57 | 1,03 |
| Map 6 | 2,79 | 81,97 | 10,92 | 1,20 |
| Map 7 | 3,13 | 83,92 | 11,48 | 1,19 |

Situation changes without using the spatial filter, 99 % of the data is unlabeled and no patter was found in the data.

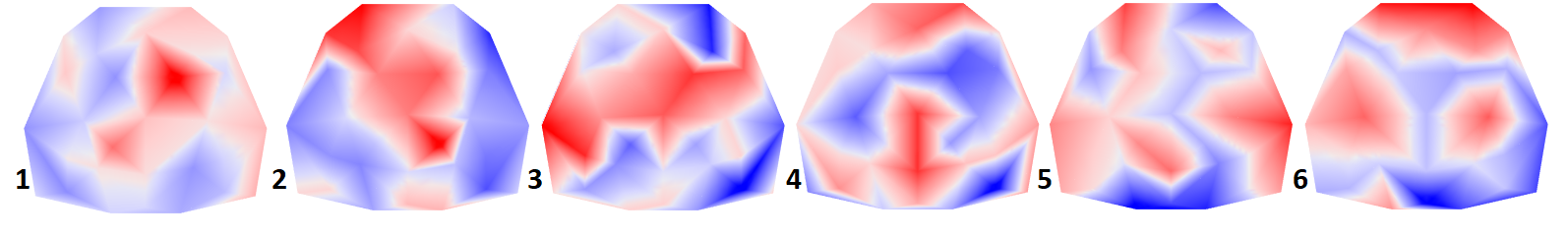

**Supplementary figure 24:** Resulting stable topographies of spatially unfiltered phantom shielded cabin EEG data recorded with 32-channel BrainProducts cap

**On the cause of induced voltage in EEG system placed in magnetic field**

According to Maxwell equations, the amount of induced voltage in closed loop placed in magnetic field, is described as follows:

$$u= \frac{d\Phi}{dt}$$

Eq. 1

Thu *u* is the induced voltage, *dΦ* is change in magnetic flux through the loop and *t* is time. The change in magnetic flux may be caused by change in loop area (*dS*) or by change in magnetic induction (*dB*) or by their interaction.

The Suppl. figure 25 shows a bit of signal acquired by EEG system placed in 3T MR bore. The sensor net was mounted on the MR spherical phantom with a conductive layer achieved by the towel soaked in saline solution. The signal was acquired while the MR machine was in “SYSTEM OFF” state and also the cooling pump and MR bore ventilation were switched off. In the following reasoning, we will consider a harmonic signal with amplitude of 10uV and frequency of either 5 or 40 Hz.

Without loss of generality, we will assume that the vector of the field is perpendicular to the vector of the loop area and the strength of magnetic induction is constant within the loop. There are two extreme situations: either 1) completely motionless loop and time-varying magnetic induction or 2) perfectly constant in time magnetic induction and time-varying area of the loop.

1)

The Eq. 1 can be:

$$A.sin\left( 2\pi ft \right)=\frac{S.d(B+g^{B}(t))}{dt}$$

Eq. 2

where the *A* and *f* is amplitude and frequency of induced signal in the loop, S is the constant loop area and B is the average magnetic induction. The g^B^(t) is a function of induction change dependent on time and it can be expressed by integrating the Eq.2 as:

$$g^{B}\left( t \right)=-\frac{A}{2\pi f.S}\cos\left( 2\pi ft \right)$$

Eq. 3

If we take rough estimation of the induced signal amplitude as above, area loop of 0.03m2 and B=3T, we get harmonic function with amplitude of 11uT or 1.3uT for the frequency of 5 or 40 Hz, resp.

2)

The Eq. 1 can be:

$$A.sin\left( 2\pi ft \right)=\frac{B.d(S+g^{S}(t))}{dt}$$

Eq. 4

where the A and *f* is amplitude and frequency of induced signal in the loop, S is the average loop area and B is the constant magnetic induction. The g^S^(t) is a function of loop area change dependent on time and it can be expressed by integrating the Eq.4 as:

$$g^{S}\left( t \right)=-\frac{A}{2\pi f.B}\cos\left( 2\pi ft \right)$$

Eq. 3

If take rough estimation of the induced signal amplitude as above and B=3T, we get harmonic function with amplitude of 0.1mm2 or 0.013mm2 for the frequency of 5 or 40 Hz, resp.

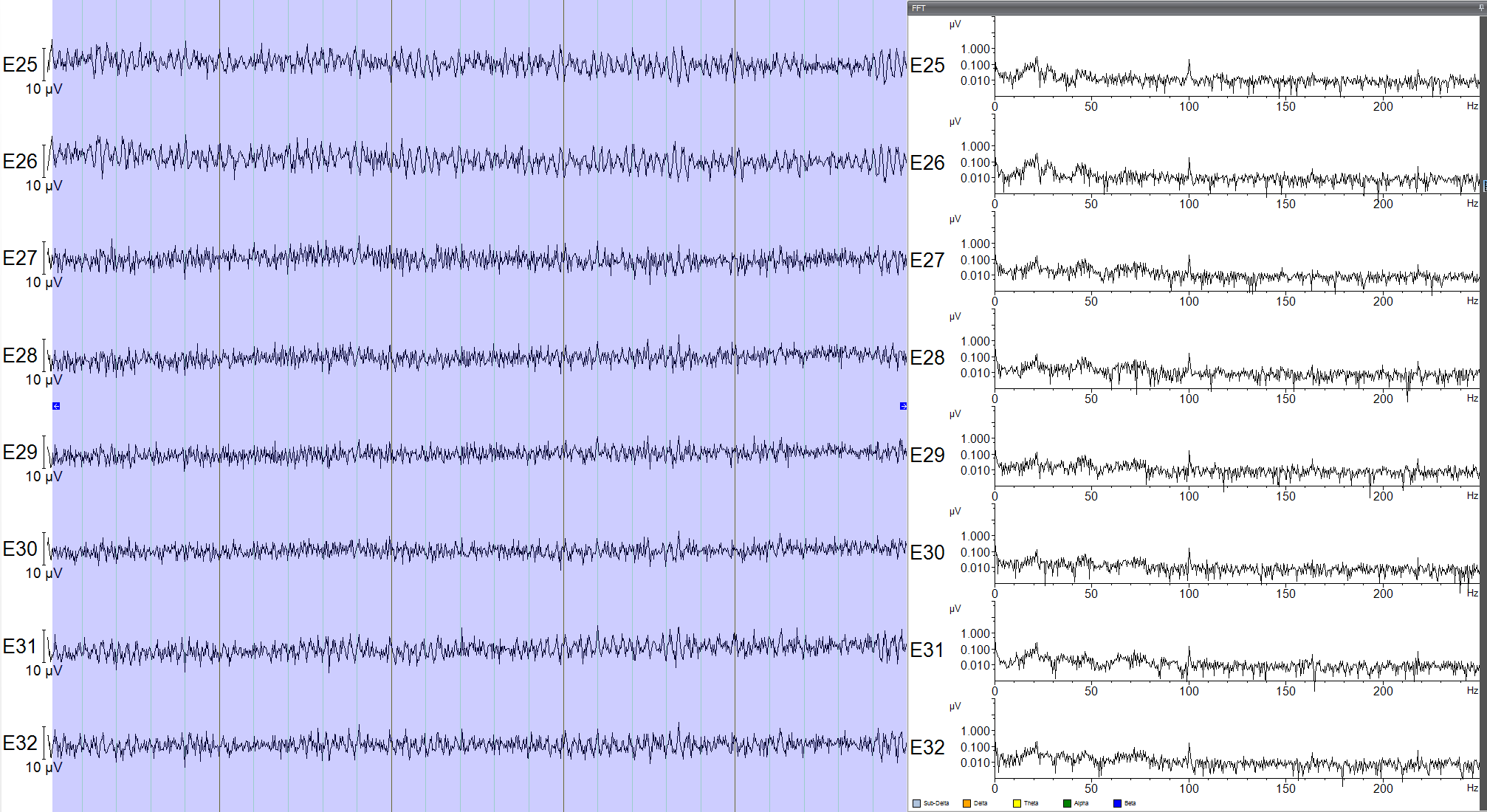

**Supplementary figure 25:** A part of phantom EEG signal acquired by EEG system placed in 3T MR bore at the left. The signal was acquired while the MR machine was in “SYSTEM OFF” state and also the cooling pump and MR bore ventilation were switched off. The amplitudes of signal reach 10uV and the majority of signal power is located between 0 and 50 Hz (see the amplitude spectrum at the right
